# The Lived Experiences of Long-Term Users of Copper-T Intrauterine Devices: A phenomenological Qualitative Study among women at Kawempe National Referral Hospital in Uganda

**DOI:** 10.64898/2026.11.04.26366407

**Authors:** Pious Igirimbabazi Ssebuddo, Jolly Beyeza-Kashesya, Imelda Namagembe, Sabinah Twesigemukama, Godfrey Aburi, Kawungu Saad Sessimba, Andrew Baluku, Kibuuka Balikudembe, Mary Mpanga, Derrick Mwanje, Khamis Musanje, Musa Sekikubo

**Author notes:** Corresponding author: - (PIS). KM and MS are joint senior authors.

## Abstract

**Background:** The Copper-T intrauterine device (Cu-T IUD) is a highly effective, non- hormonal, long-acting reversible contraceptive that can protect against pregnancy for up to 12 years, with a failure rate under 1%. Despite its clinical efficacy, early discontinuation remains a substantial challenge in Uganda, where discontinuation is approximated at 29% within the first year of use. Understanding the experiences of women using the Cu-T IUD is important to predict factors associated with discontinuation and to inform practice.

**Methods:** Between September and November 2025, we conducted twenty in-depth interviews with women of reproductive age (15-49 years) who had used a Cu-T IUD for at least two years, recruited from the family planning clinic of Kawempe National Referral Hospital, Kampala, Uganda. Data were analysed thematically, guided by the Social-Ecological Model and Lazarus and Folkman’s Transactional Model of Stress and Coping.

**Results:** Long-term Cu-T IUD users reported both positive and negative experiences shaped by individual, interpersonal, and community factors. Two themes were identified at the individual-level domain: experiences during insertion and post-insertion experiences. Negative experiences, particularly heavy menstrual bleeding, anxiety, and abdominal pain, were common and distressing. Participants employed coping strategies, including taking painkillers, using home remedies, and resting, to manage these challenges with Cu-T IUD use.

**Conclusions:** Negative experiences of Cu-T IUD are common among long-term users; women’s coping strategies such as taking painkillers lead to continued use. Client-centred pre- insertion counselling addressing both anticipated side effects and common misconceptions, alongside accessible follow-up services, may facilitate long-term Cu-T IUD use.

## Introduction

Globally, an estimated 1.1 billion women need family planning; approximately 874 million are currently using a modern contraceptive method, while 164 million have an unmet need[1]. In Sub-Saharan Africa (SSA), the overall prevalence of unmet need for family planning ranges from 23.7% to 28.8%[2–4]. In Uganda specifically, 24% of currently married women and 39% of sexually active unmarried women have an unmet need for family planning [5]. The Copper- T intrauterine device (Cu-T IUD) is a highly effective, non-hormonal contraceptive offering up to 12 years of protection with a failure rate under 1%, making it attractive to women seeking long-term solutions without frequent medical visits [6, 7]. Beyond contraception, it offers non- contraceptive benefits, including reduced risk of endometrial, cervical, and breast cancers [5, 6, 8]. Despite these advantages, utilisation of the Cu-T IUD in Uganda remains low, at approximately 2% among currently married women and 15% among sexually active unmarried women, alongside a high discontinuation rate[5]. Early discontinuation of long-acting reversible contraceptives (LARCs) remains a major challenge. In SSA, discontinuation rates within the first 12 months are notably higher: 37% in Kenya, 30% in Rwanda, 26% in Ethiopia, and 29% at KNRH[8–12]. In Uganda, nearly half (49%) of contraceptive users discontinue within two years, with IUD discontinuation rates at 22%; health concerns and side effects are the most common reasons [5, 8]. Common reasons for removal include heavy menstrual bleeding, pelvic pain, and anaemia, as well as complications such as partial expulsion[8]. [12, 13]. While existing literature extensively covers clinical efficacy and side effects of Cu-T IUD, there is a significant paucity of information regarding the lived experiences and subjective perceptions of women who Cu-T IUD as a birth control mechanism. Literature shows that the experience of usage is deeply influenced by personal factors, socio-cultural beliefs, and user interactions with healthcare providers[14]. This study aimed to engage women who had used the Cu-T IUD for more than two years to understand their experiences with this birth control method which is important to inform research and practice in Uganda. **Materials and methods**

### Study design and setting

This qualitative study used a phenomenological design, which focuses on describing the essence of a phenomenon through the perspective of those who have experienced it [15]. The study was conducted over four months (September to December 2025) at the family planning clinic of Kawempe National Referral Hospital (KNRH), Kawempe Division, Kampala, Uganda. KNRH is a specialized facility offering maternal and child health services including antenatal care, family planning (IUD/implant insertion and removal, injections, oral contraceptives, permanent methods), postnatal services, and diagnostics. The clinic operates Monday to Friday, receiving 44–58 women with Cu-T IUDs per month; approximately 12–15 have used the IUD for at least two years.

### Study population and selection criteria

The study population comprised women of reproductive age (15–49 years) who had used Cu-T IUDs continuously for at least two years and were attending KNRH for family planning services. This minimum duration was consistent with WHO recommendations for interpregnancy intervals and Uganda’s national reproductive health guidance[16]. Women unable to communicate effectively due to mental illness or other conditions affecting recall were excluded.

### Sample Size and Sampling

Twenty participants were recruited in total. Purposive sampling with a maximum variation technique was employed to incorporate diversity across age (15–24, 25–34, and 35–49 years), marital status (married/single), residence (urban/rural), occupation (formal/self-employed), education (none, primary, secondary, tertiary), and religion (Christian, Muslim, other).

Sample size was guided by the principle of information power, which proposes that adequacy is determined by richness and relevance of data rather than a fixed number[17, 18]. Phenomenological studies typically achieve sufficient information richness with 10–50 in-depth interviews (IDIs) [18]. Data collection continued until daily consensus meetings among the principal investigator (PI), research assistants (RAs), and a qualitative research expert determined that information was adequate to answer research questions. Four additional interviews were conducted after apparent data stability at the sixteenth interview to confirm no new themes were emerging, yielding a final sample of 20.

### Data collection

Data were collected using open-ended, semi-structured IDI guides developed to explore participants’ lived experiences and coping mechanisms related to long-term Cu-T IUD use. The available literature and the use of theoretical frameworks (Social-Ecological Model and Lazarus and Folkman’s Transactional Model of stress and coping) guided interview guide development, with questions structured to elicit experiences of long-term Cu-T IUD users. Participants were identified from the family planning clinic register by the clinic in-charge. Those who agreed to participate were consented and interviewed in venues providing audio and visual privacy. Interviews were co-facilitated by the PI and a female social scientist who maintained a neutral environment, took notes, and captured non-verbal cues, and interviews lasted for 45minutes. Two digital voice recorders were used simultaneously as backup. Interviews were conducted in English (n=13) and Luganda (n=7).

### Data Management

Audio files were transferred daily to a password-protected laptop. Each file received a unique participant identifier (format: IDI_(interview serial number)_(age range)). Verbatim transcription was performed within 48 hours of each interview, with 20% (four transcripts) independently reviewed by the qualitative expert for consistency. Luganda transcripts were transcribed in Luganda, translated into English by RAs, and back-translated to verify accuracy; discrepancies were resolved by consensus.

### Data Analysis

Data were analysed using a hybrid deductive-inductive thematic analysis approach [19], Guided by the SEM (Objective 1) and Lazarus and Folkman’s Transactional Model (Objective 2 ). Deductive coding was applied at the theme level, while inductive coding generated sub-themes and codes from the data. The six phases of Braun and Clarke’s approach were followed:

Phase 1: The principal investigator, RA, and qualitative research expert read through each transcript multiple times to develop familiarity with the data before coding. During this familiarisation phase, the PI read each transcript in relation to the audios, and this served as a preparatory phase for organising the data for coding. Notes were inserted in relevant sections of the transcripts to clarify the context in which statements were made, as well as to clarify statements resulting from poor sound quality.

Phase 2: Initial Coding was done manually by the PI with guidance from the expert in qualitative research, as well as the research supervisors. Printed transcripts were used as the primary working documents. Statements directly relevant to long-term Cu-T IUD use and coping strategies were extracted. These meaningful units were identified and highlighted. Coded excerpts were then manually grouped and categorised into sub-themes and themes. A master codebook was developed. This codebook was updated as new codes emerged and existing ones were refined. Independent coding of the same transcripts was done by the principal investigator and a qualitative research expert, followed by face-to-face discussion to agree on codes, sub-themes, and themes through consensus meetings.

Phase 3: Clustering Themes (Hybrid Deductive-Inductive Approach): Thematic descriptions were grouped into exhaustive categories, and from these categories, emerging sub-themes were developed.

Deductive Theme Generation: Objective 1 (Lived Experiences): Themes were pre-defined using the Socio-ecological Model, grouping data into Individual, Interpersonal, and Community levels.

Objective 2 (Coping Mechanisms): Themes were pre-defined using Lazarus and Folkman’s Model, that is, Problem-focused, Emotion-focused, and Meaning-based coping.

Inductive Sub-theme and Code Generation: Within these deductive themes, specific codes and sub-themes emerged inductively from the data. For example, “Spiritual coping” and “Cognitive reappraisal” emerged as inductive sub-themes under the deductive category of Meaning-based coping.

Phase four: Reviewing Themes: The PI and qualitative expert independently reviewed the proposed themes and their associated data extracts, grouped under each theme to assess whether they formed a coherent pattern and whether they were clearly distinguishable from other themes. Themes that contained internally inconsistent extracts were subdivided or merged with neighbouring themes through consensus meetings.

Phase five: Defining and Naming sub-themes: Attention was paid to the words, phrases, and actions of participants that captured the meaning of what they said or did. These were categorised across each theme predefined by the theoretical frameworks.

Phase Six: Producing the Report: The write-up of findings followed the structure established across phases 1–5. For each theme and sub-theme, codes and the user voices were written first. Preliminary findings were shared with 2 participants to confirm if the analysis accurately reflected their experiences. Adjustments were made based on this feedback to ensure the final report was grounded in the participants’ perspectives. The supervisors reviewed, and corrections were made during analysis, drafts of the results chapter, and provided feedback that was incorporated.

### Rigor and Trustworthiness

Trustworthiness was ensured using Lincoln and Guba’s (1985) criteria. Credibility was addressed through interviews in participants’ preferred language, open-ended questioning, prolonged engagement, probing, co-facilitation by a female social scientist, member checking (preliminary findings shared with four participants), and regular supervisor debriefs. Dependability was ensured through process documentation, and transparent application of theoretical frameworks. Confirmability was maintained through dual independent coding. Transferability was enhanced through maximum variation and a detailed description of the research process; reporting of findings and limitations were documented to enable readers to assess the applicability of these research findings to their own settings..

### Researcher Reflexivity

The Lead investigator, a male resident in Obstetrics and Gynaecology at KNRH, recognized potential risks of social desirability and confirmation bias. Mitigation strategies included co-facilitation by a female social scientist, independent dual coding, member checking, and consensus-based analysis with a qualitative expert.

### Ethical Approval

Ethical approval was obtained from Makerere University School of Medicine Research and Ethics Committee (Mak-SOMREC-2025-2110) and administrative clearance from KNRH. All participants provided informed written consent. Confidentiality was maintained through participant numbering; no identifying information was included in transcripts or reports. Two participants withdrew voluntarily without implications for their care.

## Results

Most participants were aged 25-34 years, married, had 3-5 children, and were self-employed, as shown in table 1 bellow.

**Table 1.** Sociodemographic characteristics of long-term users of Cu-T IUDs at KNRH (n=20)

| **Characteristic** | **Category** | **Frequency (n=20)** |
|---|---|---|
| Age category | 15-24 years | 5 |
|  | 25-34 years | 8 |
|  | 35-49 years | 7 |
| Marital status | Single | 6 |
|  | Married | 14 |
| Residence | Urban | 15 |
|  | Rural | 5 |
| Religion | Anglican | 6 |
|  | Catholic | 2 |
|  | Muslim | 5 |
|  | Pentecostal | 5 |
|  | Other | 2 |
| Education level | No formal education | 3 |
|  | Primary | 9 |
|  | Secondary | 5 |
|  | Tertiary | 3 |
| Occupation | Formal employment | 7 |
|  | Self-employed | 13 |
| Number of children | ≤2 | 7 |
|  | 3-5 | 10 |
|  | >5 | 3 |

### Lived experiences of long-term users of Cu-T IUDs

Participants described a range of positive and negative experiences associated with long-term Cu-T IUD use. These experiences were shaped by factors across domains at the Social-Ecological Model (SEM) as presented in Table 2 below.

**Table 2.** Themes, sub-themes, and codes identified for the lived experiences of long-term Cu-T IUD users at KNRH.

| **Themes** | Sub themes | **Representative codes** |
|---|---|---|
| **Individual level domain** |  |  |
| Experience during insertion | Physical experiences following insertion | Post-insertion bleeding; post-insertion abdominal pain; absence of physical side effects |
|  | Anxiety during the insertion process | Fear of the insertion procedure |
| Post-insertion experience | Menstrual cycle changes | Heavy menstrual bleeding; normal menstrual cycles |
|  | Emotional and psychological experiences | Fear of unintended pregnancy; anxiety about the IUD changing position |
|  | Sexual experiences | Increased sexual confidence; increased sexual desire |
| Interpersonal-level domain |  |  |
| Relationship encounters | Anxiety or reassurance arising from partner influence | Anxiety arising from partner concerns about sexual effects; partner support leading to reassurance |
|  | Anxiety or confidence arising from peer and family influence | Confidence arising from relatives and peer-shared positive experiences; Anxiety arising from peer-shared negative experiences |
| Community Level domain |  |  |
| Participants experiences with hospital interactions | Reassurance from follow-up visits and continuity of care | Difficulty locating IUD strings; imaging to confirm placement; scheduled follow-up reviews |
| Anxiety arising from myths and misconceptions in the community | Living with anxiety associated with rumoured side-effects | Anxiety arising from myths in the community linking IUD to cancer/fibroids; device migration; fears of infertility/infection |
|  | Anxiety arising from past negative experiences observed in the community | Fear of IUD failure; increased fear and anxiety of complications |

### Lived experiences of copper T IUD users at individual-level domain

Two themes emerged: Experiences during insertion and post-insertion experiences. Experiences during insertion included physical experiences following insertion; Post-insertion experiences included menstrual cycle changes, emotional and psychological experiences, sexual experiences, and decision-making experiences.

### Experiences of long-term Cu-T IUD users during insertion

Physical experiences following insertion varied; some women described post-insertion bleeding and abdominal pain that diminished over time and was managed with analgesics, while others reported no notable side effects.

> *“I got blood spotting for the first 3-4 days, then even my first periods were heavier than the usual ones for the first 5 months … I would get uncomfortable, which made it hard to go to work some days… but later I got used to putting big cotton pads that would last longer” (IDI_15_30-34yrs)*.
>
> *“Within the first 5 months of IUD usage, I experienced abdominal cramps, but they were bearable after taking painkillers, and they did not disturb my day-to- day activities” (IDI_05_25-29yrs)*.

Most participants experienced fear and anxiety at the insertion of the IUD however, this was reduced by pre-insertion counselling and provider reassurance.

> *“I became anxious, and I told the nurse that I was scared, and she tried to calm me down, . . . I prepared for the worst scenario of bleeding and abdominal pain, but it never happened. “ (IDI_09_35-39yrs)*
>
> *“The nurse* knew *that I was afraid during and after the insertion, and so she kept asking how I felt, and I opened up to her about how I was feeling crampy abdominal pain, and I was scared of it penetrating my uterus. She counselled me and gave me painkillers which helped me to keep on using the IUD” (IDI_02_30-34yrs )*

### Post-insertion experiences of long-term Cu-T IUD users

The majority of participants experienced menstrual changes, especially heavy bleeding, and some developed contemplation about removing it.

> *“In the* first *3 months, I got a lot of bleeding that made me worried, couldn’t go to work, and I even thought I was going to die, so I went to the hospital to have my IUD removed, but after checking my blood, the doctor advised me to keep it and after completing treatment, the bleeding stopped” (IDI_04_40-44yrs)*

Some participants faced emotional and psychological experiences centred on fear of unintended pregnancy, anxiety about the IUD changing position, and fear of the insertion procedure itself, often resolved through reassurance and clinical confirmation of correct placement:

> *“My* biggest *fear was for the IUD to move due to heavy bleeding, and then I get pregnant*
>
> *. . . I have done three scans that show that it is placed in the uterus and no pregnancy*.
>
> *. I now feel* comfortable *after using it for 4 years” (IDI_04_40-44yrs)*
>
> *“I used to* insert *my fingers down to feel for the threads when I was bathing, and was always scared… the health workers actually told me that I should stop doing it… Whenever I came for review I would do a scan and the IUD was in the uterus… Am now confident I am no longer worried” (IDI_16_30-34yrs)*.

Sexual experiences following Cu-T IUD use were varied; some participants reported change in sexual appetite due to increased sexual desire associated with increased lubrication.

> *“Due to increased vaginal fluids I started getting while using the IUD, the desire for sex increased more than usual, and many times my husband realized the change… and my level of satisfaction increased, which worsened during ovulation… I was encouraged to continue using it” (IDI_09_35-39yrs)*.

### Lived experiences of long-term Cu-T IUD users at Interpersonal-level domain

Participants described how partners, peers, and family members directly shaped their experience of long-term use. Caution from friends about perceived changes during intercourse led to anxiety feelings, while partner support and reassurance facilitated continued use.

Peer and family influence operated through both encouragement and cautionary advice, and through the sharing of positive or negative experiences, participants experienced encouragement to continue using Cu-T IUD and anxiety and fear, respectively.

> *“When my friends at work who had used the method told me things about feeling abdominal pain, and increased bleeding . . . it increased my fear . . . However, when I reached the hospital, I found and shared with a woman who had used it for 10 years .* .
>
> *. I* got *encouraged and kept using the method“* (*IDI_10_35-39yrs)*

Some participants described normalizing usage of the IUD based on receiving supportive and reassuring feedback from their partners. Partner support often facilitated continued use of the IUD and increased confidence in the method.

> *“I told him that I removed the implant and inserted the IUD. He encouraged me to continue using it, which made me feel that he understood me, and I persevered with side effects until my body got used to it”* (*IDI_09_35-39yrs)*

Interactions with peers and family members also shaped participants’ experiences of Cu-T IUD use. Advice from relatives and peer-shared positive experiences played an important role in normalizing usage, thus shaping users’ lived experiences and decision to continue using Cu T-IUD.

> *“My grandmother is a nurse, and she recommended that I should use the IUD, and she kept on encouraging me whenever I got abdominal pain. But now I have a serious boyfriend, and I want to get pregnant.” (IDI_05_15-19yrs)*
>
> *“There is a woman I found in the hospital who had been using the IUD. She told me that the IUD was her best method for family planning . . . no daily monitoring required, no pregnancy worries, and she was satisfied with the method . . . This increased my trust in the method and continued using it” (IDI_10_35-39yrs)*

Conversely, participants’ interaction with peers who had removed IUDs due to side effects reinforced fears about the Cu-T IUD users. Hearing about negative experiences from others contributed to anxiety, uncertainty, and, for some women, led them to consider discontinuing the method.

> *“When my friends at work who had used the method told me things about feeling abdominal pain, and increased bleeding they experienced that stopped after removal* .
>
> *. . it increased my fear, I started feeling anxious and decided to remove it . . . However, when I reached the hospital, I found and shared with a woman who had used it for 10 years . . . I got encouraged and kept using the method” (IDI_10_35-39yrs)*

### Lived experiences of long-term of copper T IUD users at the community level. Interaction with community members

Participants described how community narratives and myths generated lived emotional experiences. Beliefs linking the IUD to cancer, fibroids, device migration, infertility, and infection were frequently encountered and generated fear, anxiety, and self-doubt, sometimes prompting unplanned clinic visits to seek reassurance. Witnessing negative outcomes among community members using IUDs such as device expulsion, heightened these fears among participants, yet most of them described continuing use despite these community-level influences:

> *“People at my workplace keep saying that if you spend a long time with the IUD, it can make you get fibroids and it can disappear in your body. This made me fear for my health, and I came today for a review . . . The scan I did has shown that I don’t have fibroids and the IUD is in the right place.” (IDI_16_30-34yrs)*
>
> *“In a village meeting, I overheard people talk ill of the IUD as they say it gives people cervical cancer, and that is why I was very scared because I want to give birth before I get problems” (IDI_09_35-39yrs)*
>
> *“My workmate told me that the IUD weakens the uterus, and then you will always get miscarriages. I first removed it after use for one year. I conceived; the pregnancy and child were okay. Now I used the IUD again, and I have kept it for now 3 years, and it has worked for me” (IDI_06_25-29yrs)*
>
> *“People say that it causes a lot of abnormal discharges, and anytime you go to urinate, you can get infections because after they insert the IUD, you remain open down there (cervix), and I used to get frequent, increased vaginal discharge ever since I put it . . . but the doctor told me that it’s not an infection but increased vaginal fluids. I got used to it, and sometimes I use a pad. “ (IDI_09_35-3yrs)*
>
> *“At the saloon, people were saying that an IUD can cause infertility . . . I used it for 3years at, first removed it, and I conceived immediately . . . this is my second time to use it, and I will use it again after delivering the third child” (IDI_02_30-34yrs)*

### Interaction with hospital environment

Participants described continuity of care, including scheduled follow-up and access to ultrasound imaging to confirm correct device placement; this provided reassurance, although difficulty locating IUD strings during examination and the unplanned cost of imaging were sources of stress and financial burden for some participants:

> *“I wanted to remove the IUD and conceive, but I had a problem. The doctor told me she is not seeing it, so they sent me for a scan, and I had to look for money and pay . . . the scan showed that the IUD is in the right place” (IDI_06_25-29yrs)*
>
> *“When I got back to the hospital, I did the scan to see the position of the IUD. It was well placed. I felt confident again and continued using the IUD, especially when I knew that they could see what was happening inside the uterus, so this kept me going, and I kept on doing checkups” (IDI_14_40-44yrs)*

### Coping strategies users employ to manage negative experiences of Cu-T IUD

The major negative experiences identified were vaginal bleeding, increased vaginal discharge, abdominal pain, and fears and anxiety related to misconceptions and procedural concerns.

This section captures the various coping strategies participants employed to manage the physical, emotional and social challenges associated with long-term Cu-T IUD use. These were categorized according to Lazarus and Folkman’s model into problem-focused, emotion-focused, and meaning-based coping strategies. The themes and codes identified for each coping domain are summarized in Table 3 below.

**Table 3.** Coping strategies used to deal with negative experiences associated with Long-Term use of Cu-T IUD.

| Theme | Sub-theme | Negative Experiences | Codes |
|---|---|---|---|
| Problem-focused strategies | Symptom Containment Practices | ➢ Vaginal bleeding/ Increased vaginal fluids | ➢ Using stronger pads and Pampers<br>➢ Changing pads more often |
|  |  | ➢ Abdominal pain | ➢ Taking painkillers<br>➢ Using home remedies<br>➢ Resting and limiting activity |
|  | Health-seeking behaviors | ➢ Anxiety and fears<br>➢ abdominal pain<br>➢ Vaginal bleeding | ➢ Went back to the hospital |
|  |  | ➢ Abdominal pain<br>➢ Vaginal bleeding | ➢ Use of prescribed treatment |
|  |  | ➢ Fears and anxiety<br>➢ Abdominal pain | ➢ Provider counselling and reassurance |
|  |  | ➢ Fears and anxiety<br>➢ Abdominal pain | ➢ Doing abdominal scan to alley anxiety |
| Emotion-Focused strategies | Emotional self-regulation | ➢ Anxiety and fears | ➢ Positive self-talk |
|  |  | ➢ Fears and anxiety | ➢ Endurance and tolerance |
|  | Social support from others | ➢ Fears and anxiety | ➢ Partner reassurance |
|  |  | ➢ Fears and anxiety<br>➢ Abdominal pain | ➢ Emotional comfort from relatives |
|  |  | ➢ Increased vaginal fluids | ➢ Reassurance from positive stories |
| Faith and making meaning | Reliance on faith | ➢ Anxiety and fears | ➢ Praying about it |
|  |  | ➢ Anxiety and fears<br>➢ Abdominal pain | ➢ Trusting God to help |

#### 4.3.1 Problem-Focused strategies

Participants adopted practical, symptom-containment measures, including switching to more absorbent sanitary materials, more frequent pad changes, use of analgesics, home remedies such as ginger tea, and rest during episodes of heavy bleeding or pain.

> *“I used to use pads but, switched to pampers because my periods were heavy . . . and it worked for me until my periods stabilised” (IDI_03_30-34yrs)*
>
> *“I started to change my pads frequently and using towels more often due to increased vaginal fluids . . . When bleeding increased, I had to bear with changing the towels until my body got used to it” (IDI_02_30-34yrs)*

Health-seeking behaviours included returning to the hospital for review, adherence to prescribed treatment, seeking provider counselling and reassurance, and undergoing abdominal ultrasound to confirm device placement and allay anxiety:

> *“Within the first 5 months of IUD usage, I experienced serious cramps, but they were bearable after taking painkillers. You take some painkillers to reduce the pain, and you are free to work.” (IDI_05_25-29yrs)*
>
> *“To deal with the pain I would drink warm ginger tea and rest a lot even at the shop during menstrual periods due to increased abdominal pain, and this helped me until my period became regular.” (IDI_0 3_30-34yrs)*

Several participants reported attending follow-up visits at KNRH when experiencing unusual abdominal pain, prolonged bleeding, or fears and anxiety. Regular medical reviews provided reassurance, timely management of symptoms, and guidance for continued use.

> *“I came back to seek medical help after getting excessive bleeding, and the health workers examined me, gave me treatment, clearly explained my situation, and they comforted me so well . . . so I was encouraged to continue using the IUD” (IDI_04_40-44yrs)*
>
> *“People at my workplace keep saying that if you spend a long time with it, it can make you get fibroids and it can disappear . . . this makes me fear for my health, and I came today for a review . . . The scan I did has shown that I dont have fibroids and its in the right place . . .” (IDI_16_30-34yrs)*

Some participants reported receiving counselling and reassurance by health workers and this helped in alleviating fears related to Cu-T IUD penetrating the uterus and abdominal pain.

> *“The nurse knew that I was afraid, during the review visit and so she kept asking how I felt, and I opened up to her how I was feeling crampy abdominal pain, and I was scared of it penetrating my uterus. She counselled me and gave me pain killers which helped me to keep on using the IUD” (IDI_02_30-34yrs)*
>
> *“I went for the checkup after one month, and I did a scan to see if the IUD was inserted perfectly, and so it was. This helped to trust the health worker messages, and I kept on using the IUD” (IDI_09_35-39yrs)*

### Emotion-Focused Strategies

Participants used positive self-talk and endurance to regulate emotional responses to discomfort, and drew on social support from partners, family, and peers for reassurance and emotional comfort:

> *“I know that I am a mother, and this is my responsibility . . . So I kept on telling myself that I could make it if others had made it. This helped to keep on focusing on the positives of the IUD rather than the stressful negative stories in the community, and the abdominal pain after a few months, my body adjusted” (IDI_13_25-29yrs)*
>
> *“Whenever I was worried about what might happen or the side effects of the IUD, my husband would comfort me, and since he also supported the decision, it somehow helped me feel less anxious and confident, and I continued using the method” (IDI_07_20-24yrs)*
>
> *“My mother shared with me her experience with family planning. She also used the IUD, and she told me not to have any worries, so I felt it was a normal thing and continued using the IUD despite mild abdominal pains.” (IDI_11_35-39yrs)*

### Faith and meaning-based strategies

Participants drew on prayer and trust in God to find strength and resilience during periods of discomfort, and used cognitive reappraisal, accepting provider explanations of side effects as normal and temporary, to sustain continued use:

> *“At first, I was worried because of the abdominal pain and increased bleeding, but later I accepted that it was part of the body adjusting. I told myself it would not last forever, and with time, it reduced” (IDI_07_20-24yrs)*
>
> *“Sometimes I would pray when I used to get abdominal pain, and I got strength to move on because God can help, and since I am a believer, I knew that there would be a change, and this helped me a lot” (IDI_08_25-29yrs)*
>
> *“I said bismillah and left everything to God. When using medication for the abdominal pain, I kept on trusting God . . . I had a lot of fears, and I was sorted with time” (IDI_16_30-34yrs)*

## Discussion

This study set out to explore the lived experiences of long-term Cu-T IUD users across the domains of the Social-Ecological Model (Objective 1) and to describe the coping strategies participants used to manage these experiences, guided by Lazarus and Folkman’s Transactional Model of Stress and Coping (Objective 2). Both objectives were achieved: participants’ accounts captured experiences shaped by an interplay of individual, interpersonal, and community-level factors. Two themes were identified at the individual-level domain: experiences during insertion and post-insertion experiences

The most striking finding was that participants sustained long-term use despite frequently distressing experiences, including heavy menstrual bleeding, abdominal pain, and fear generated by community myths linking the Cu-T IUD to cancer, infertility, and device migration. Continuation therefore did not depend on the absence of adverse experiences but on the availability of counselling, reassurance, and social and spiritual resources that helped women reinterpret and tolerate them as temporary and manageable. This is consistent with evidence from Nigeria showing that IUD side-effects diminish substantially within the first 6– 12 months of continued use [2], and with WHO guidance identifying pre-insertion counselling, rather than the absence of side effects, as the pivotal enabler of continuation [20].

Most participants experienced heavy menstrual bleeding and abdominal pain and were often distressing, yet participants persisted with use. This is consistent with the study done in Nigeria by Ayogu et al that demonstrated that side-effects of the IUD decreases significantly over time among continuing users, with bleeding typically stabilizing within 6–12 months[2]. WHO’s Selected Practice Recommendations for Contraceptive Use similarly emphasize pre-insertion counselling as a standard of care, enabling women to persist through the adaptation period [20]. Sexual experiences were varied. Some participants experienced increased sexual appetite due to increased sexual desire, which was linked to enhanced lubrication following Cu-T IUD use. Partner sexual concerns, including perceptions of changes in vaginal secretions, were a source of relationship tension and anxiety among users of the Cu-T IUD. This was consistent with the study findings of male partner anxiety as a barrier to IUD uptake in Uganda[21].

Most participants experienced anxiety during Cu-T IUD insertion however health providers’ counselling became a determinant of method continuation, particularly among women with prior adverse hormonal contraceptive experiences. This is consistent with a study done in the USA by Habacher and Kavanaugh that demonstrated that IUD pre – insertion counselling on how a method works is associated with higher satisfaction and continuation [22]. In SSA, pre-insertion counselling has similarly been associated with sustained LARC use [9, 12].

Fear driven by community myths and misconceptions about IUD usage emerged as the most consistent participants’ experience, arguably more so than the physical side-effects themselves. Some participants explained how they lived with fear and anxiety due to myths and misconceptions in the community linking IUDs to cancer, infertility, device migration, and infections, and echoed findings from other Ugandan settings, including Luweero and Entebbe [21] as well as broader Sub-Saharan Africa linking such misconceptions to early removal[8, 12, 23]. That these fears persisted even among women already committed to long-term use of the IUD suggests that myth-related anxiety among Cu-T IUD users functions more as a recurring burden that continuing users must actively manage.

Turning to Objective 2, participants’ own coping strategies were central to how they coped with these negative experiences with long-term Cu-T IUD use. Problem-focused strategies, including switching to more absorbent sanitary materials and using analgesics, reflect adaptive resourcefulness documented among women managing heavy menstrual bleeding in low-income urban settings, including Kampala [24]. NSAID use for pain management is consistent with WHO recommendations and Cochrane evidence supporting mefenamic acid and ibuprofen for IUD-associated abdominal pain and blood loss reduction [25].

Emotion-focused strategies, particularly positive self-talk and partner reassurance, are consistent with [26] finding that women who sustained IUD use commonly described a reorientation toward contraceptive benefit over immediate discomfort.

Meaning-based coping through prayer and cognitive reappraisal reflects the centrality of faith in health behaviour in Uganda’s predominantly Christian and Muslim population [27]. Provider-facilitated cognitive reappraisal, understanding side effects as normal and temporary, was a critical mediating factor in sustained use, consistent with evidence from Sweden [15] and SSA [28].

## Limitations of the study

Participants were recruited at KNRH, a national referral hospital with specialist care and diagnostic equipment; findings may not be representative of long-term IUD users at lower-level health facilities across Uganda. Social desirability bias was mitigated through co-facilitation by a female social scientist and use of probing techniques. The study captured only the perspectives of women who persisted with the method; the experiences of early discontinuers were not explored. Partner perspectives were accessed solely through women’s accounts; future research should include partner-inclusive designs.

## Conclusions

Women’s experiences with long-term Cu-T IUD use encompass both positive and negative dimensions, shaped by interactions across individual, interpersonal and community levels. Two themes were identified at the individual-level domain: experiences during insertion and post-insertion experiences. Negative experiences, particularly heavy menstrual bleeding, abdominal pain, and fear generated by community myths, were common and often distressing.

Coping strategies by IUD users include adherence to provider guidance, family and partner social support, enabling women to normalize side effects and sustain long-term use. These findings underscore the importance of comprehensive, client-centred pre-insertion counselling that addresses anticipated side effects, corrects myths and misconceptions, engages partners where appropriate, and provides emotional reassurance.

Future qualitative research should explore the experiences of women who discontinue Cu-T IUD use early, as this study captured only the perspectives of women who persisted with using the method, and should incorporate partner perspectives directly, as partner influence emerged as a significant theme accessed in this study only through women’s reports.

## Data Availability

All data underlying this study consist of in-depth interview transcripts containing sensitive personal health information shared by participants under assurances of confidentiality. In accordance with PLOS ONE's policy for qualitative research data, representative transcript excerpts supporting all reported themes are included within the paper. Full transcripts cannot be shared publicly, as doing so would violate participant consent agreements and risk identification of individuals who disclosed stigmatised health experiences. Anonymised thematic data are available from the corresponding author on reasonable request, subject to approval from the Makerere University School of Medicine Research and Ethics Committee (SOMREC; approval reference: Mak SOMREC-2025-2110) and compliance with applicable data protection requirements.

## List of abbreviations

Cu-T IUD: Copper-T Intrauterine Device
IDI: In-depth Interview
KNRH: Kawempe National Referral Hospital
LARC: Long-Acting Reversible Contraceptive
PI: Principal Investigator
RA: Research Assistant
SEM: Social-Ecological Model
SOMREC: School of Medicine Research and Ethics Committee
SSA: Sub-Saharan Africa
UDHS: Uganda Demographic and Health Survey
WHO: World Health Organization.

## Consent for publication

Not applicable.

## Availability of data and materials

The datasets generated and/or analysed during the current study, including de-identified interview transcripts, are available from the corresponding author on reasonable request, subject to ethical approval for secondary use, given the qualitative and potentially identifying nature of the data.

## Authors’ contributions

All authors read and approved the final manuscript.

**PIS** conceived and designed the study, collected and analysed the data, interpreted the results, and drafted the manuscript.

**JBK, KM** and **IN** contributed to refining the research concept and methodology, provided feedback on manuscript drafts, supervisory roles and ensured adherence to ethical standards throughout the research process.

**GA, ST, SSK, MM, DM** and **KB** participated in writing and refining the initial drafts of the manuscript, reviewed and provided feedback that was used to refine the initial drafts of the manuscript, contributed to reviewing and provided feedback towards refining the final manuscript drafts.

**MS** gave overall oversight and supervision, contributed to the study design and methodology, supervised data analysis, and critically reviewed the manuscript for scientific content.

## Acknowledgments

The authors thank the women who participated in this study and shared their experiences, the staff of the Kawempe National Referral Hospital family planning clinic, and the research assistants who supported data collection and transcription.

## Authors’ information

Not applicable.

## APPENDIX I INFORMED CONSENT

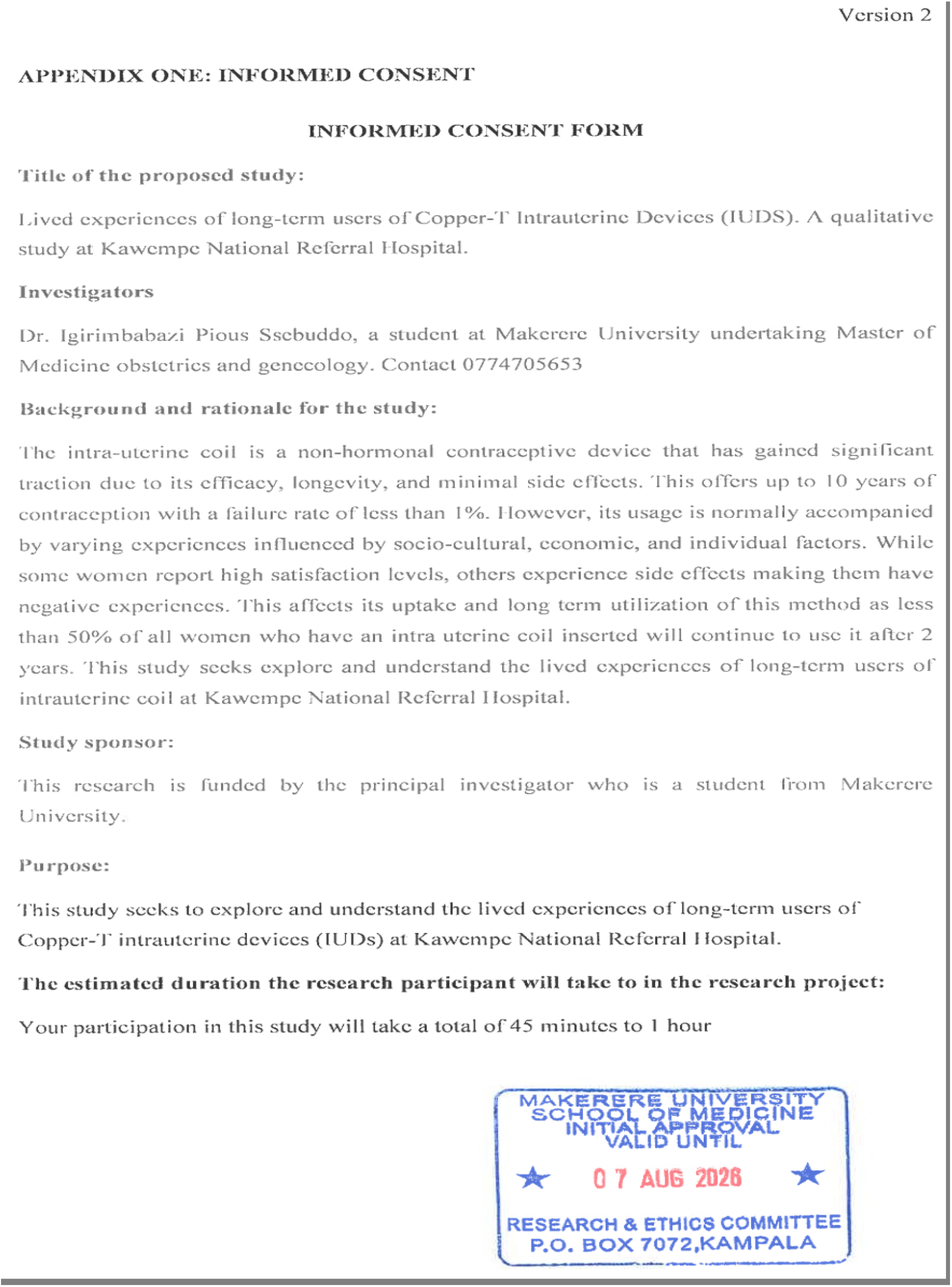
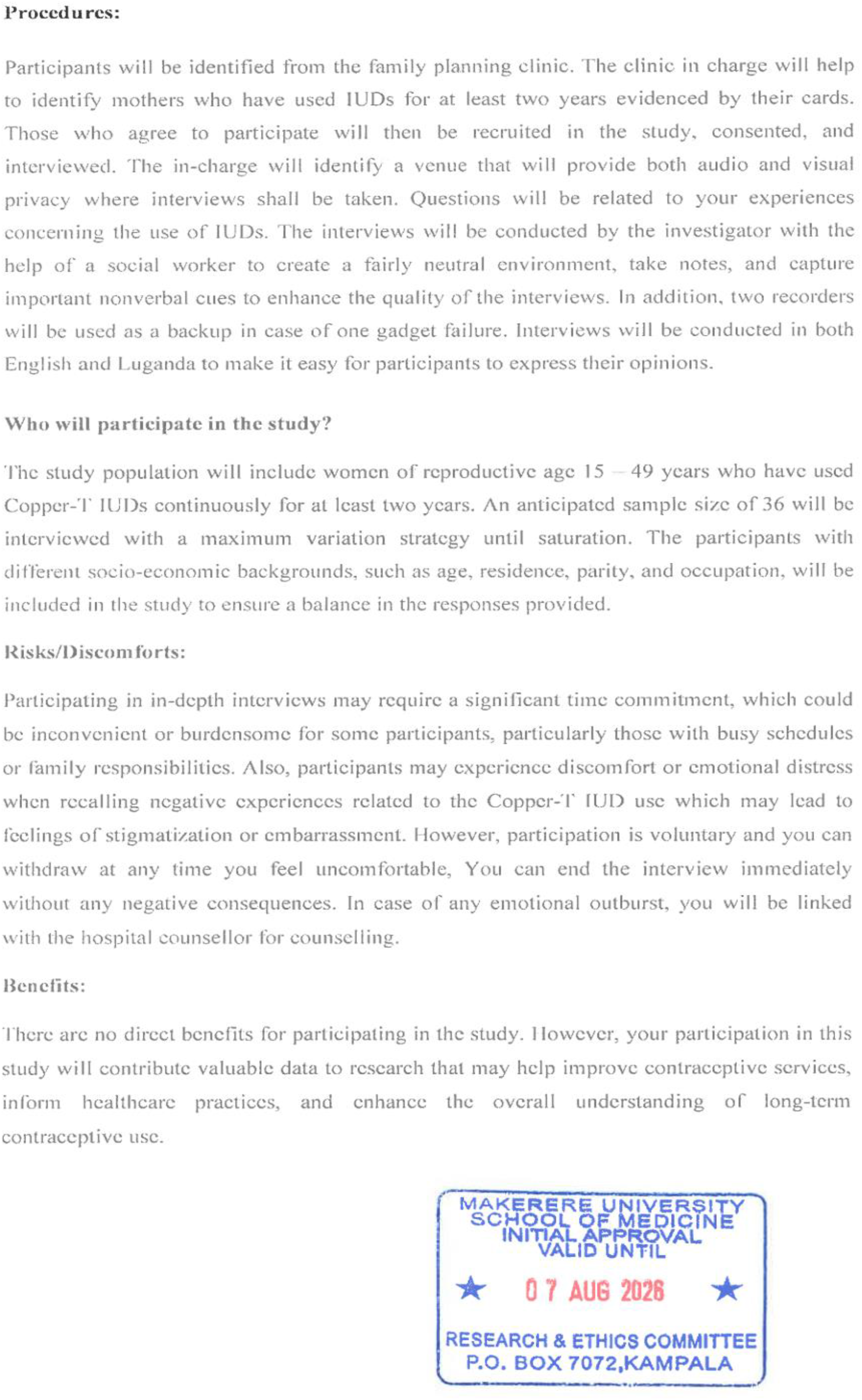
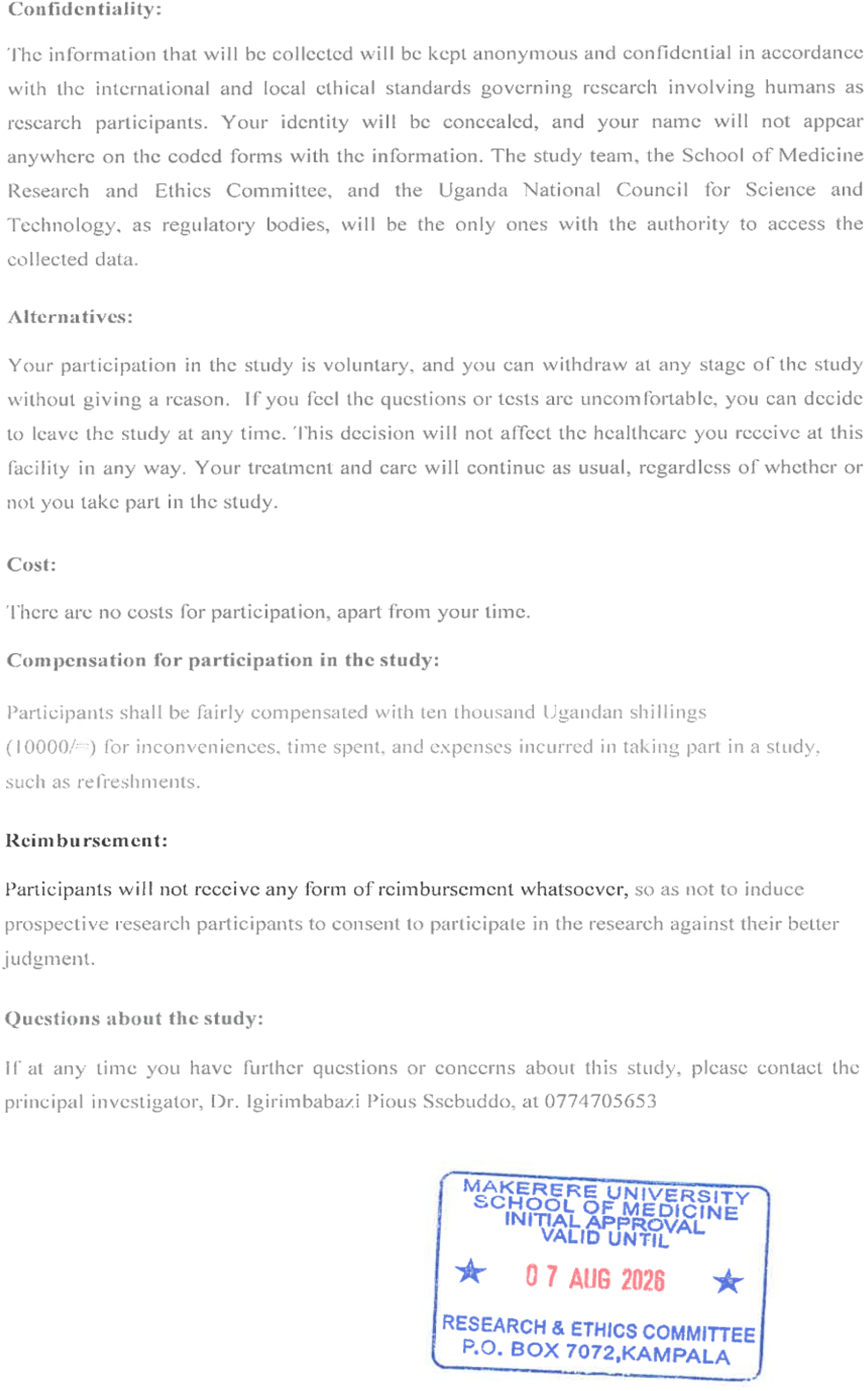
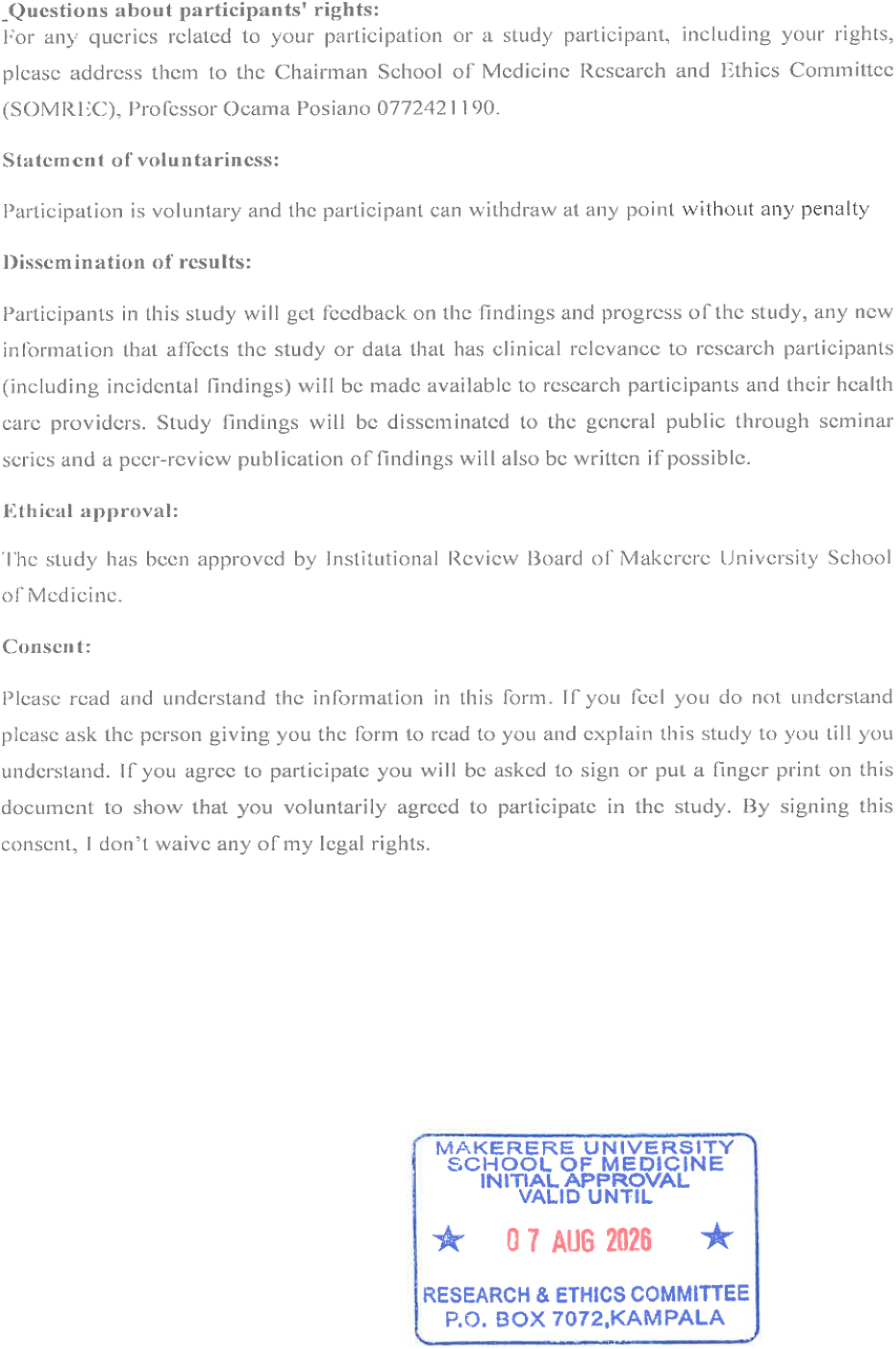
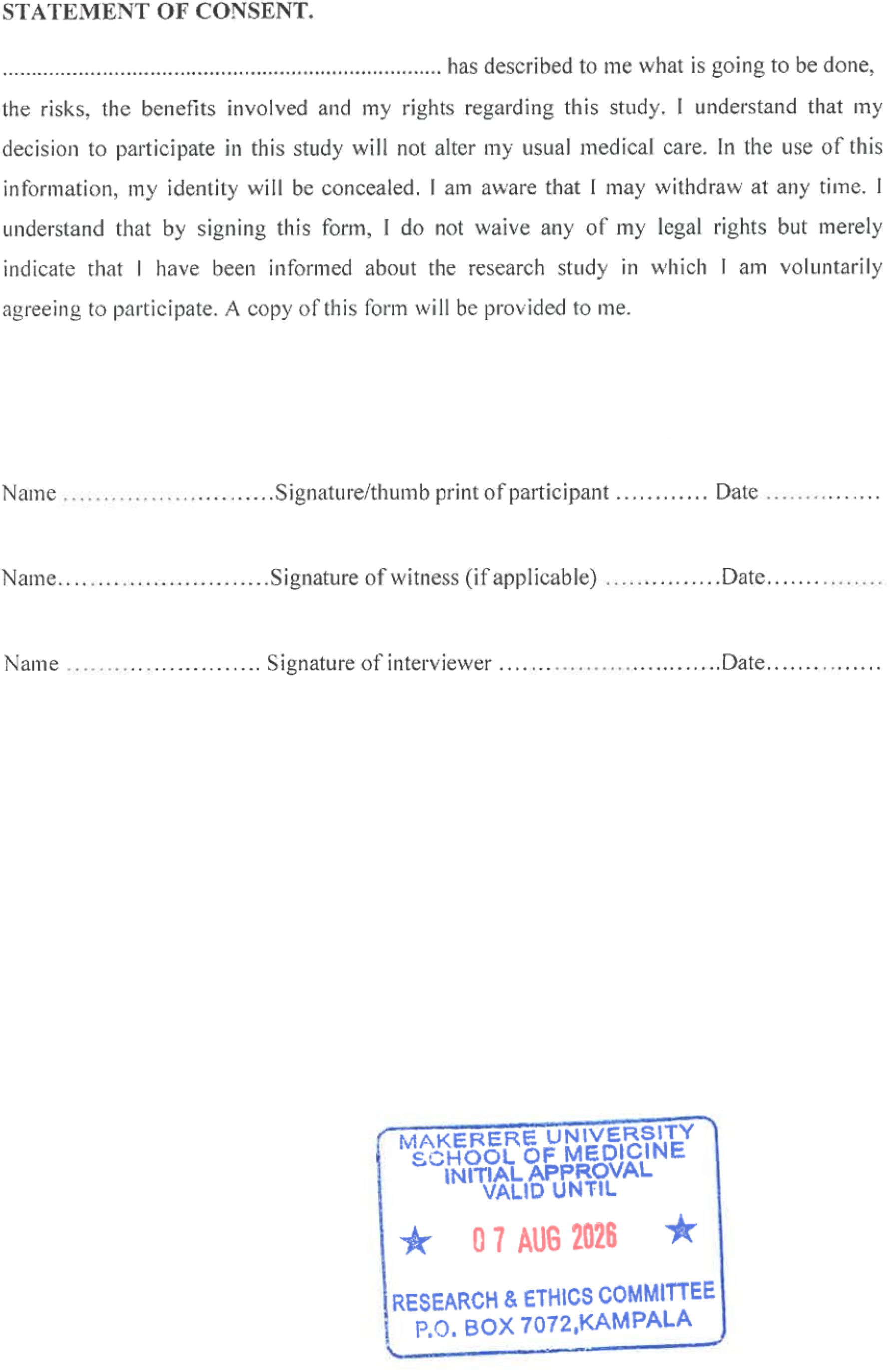

## APPENDIX II INTERVIEW GUIDE

Hello, my name is . I am conducting a Study on the **Lived Experiences of Long-Term Users of copper-T Intrauterine Devices (IUDs**). A Qualitative Study at Kawempe National Referral Hospital. Your participation in this study will contribute valuable data to research that may help improve contraceptive services, inform healthcare practices, and enhance the overall understanding of long-term contraceptive use. We appreciate your experiences, opinions, and suggestions for ways to improve family planning service delivery in Uganda. I am requesting that we audio-tape this conversation so that we capture all your contributions. I do not want to lose some of your valuable ideas. After transcribing the information, we will destroy the audio recordings. **There are no right or wrong answers; therefore, feel free to give us what you think.**

**Section 1: Demographic Information**

Before we begin, could you provide some basic background information?

1. Age:
2. Marital status:
3. Religious affiliation
4. Number of children:
5. Educational level:
6. What is your occupation?

**Introduction.**

1. What is your understanding of the Copper-T IUD?
2. Tell me how you decided to use the IUD. What led you to choose it?
3. Probe for:
4. Whether they discussed with family and/or friends
5. where the Cu-T IUD was inserted from.
6. How long have you been using the Copper-T IUD?

**Section 2: Experiences with Copper-T IUD**

1. What is your experiences about the process of insertion.
2. Tell me about your overall experiences of using the Copper-T IUD? Probe for;

- What benefits have you experienced from using the Copper-T IUD?
- How has the Copper-T IUD impacted your daily life or routines, if at all?
- In your opinion, how has the spouse shaped your experience with long-term IUD usage?
- In your opinion how have your immediate family and friends shaped your long-term IUD usage
- In your opinion how has your community impacted your long-term IUD usage?
3. In your opinion what is the level of satisfaction that you obtained with using Copper-T IUD in preventing pregnancy?
4. In your opinion what positive aspects does the spouse say about the Copper-T IUD in preventing pregnancy?
5. In your opinion what do your immediate family and friends say about the use of Copper-T IUD in preventing pregnancy?
6. Please describe any challenges you’ve experienced while using the Copper-T IUD? (Probe for personal, partner challenges)

Probe for:

- Tell me what has changed since you started using IUD. (These may include *Side effects, discomfort, emotional issues), Probe for how for each item mentioned*.
- Menstrual changes: How has your menstrual cycle changed since you started using IUD?
- Sexual changes: How has the use of IUD affected your sexual life?

8. What are the coping strategies you used to overcome these challenges above?

9. How have these experiences affected your decision to continue or stop using the IUD?

10. What other factors have influenced your decision to continue using the Copper-T IUD for this long?

Probe: How do factors like family planning needs, family planning policies, societal norms, or healthcare advice play a role?

11. Based on your experience, what advice would you give to other women who are considering uptake of Family planning?

12. What suggestions do you have for improving the support provided to Copper-T IUD users at Kawempe National Referral Hospital?

13. Is there anything else you would like to share about your experience with the Copper-T

IUD that we haven’t discussed?

Thank you so much for sharing your experiences with us.

## APPENDIX III COMMUNITY ENGAGEMENT

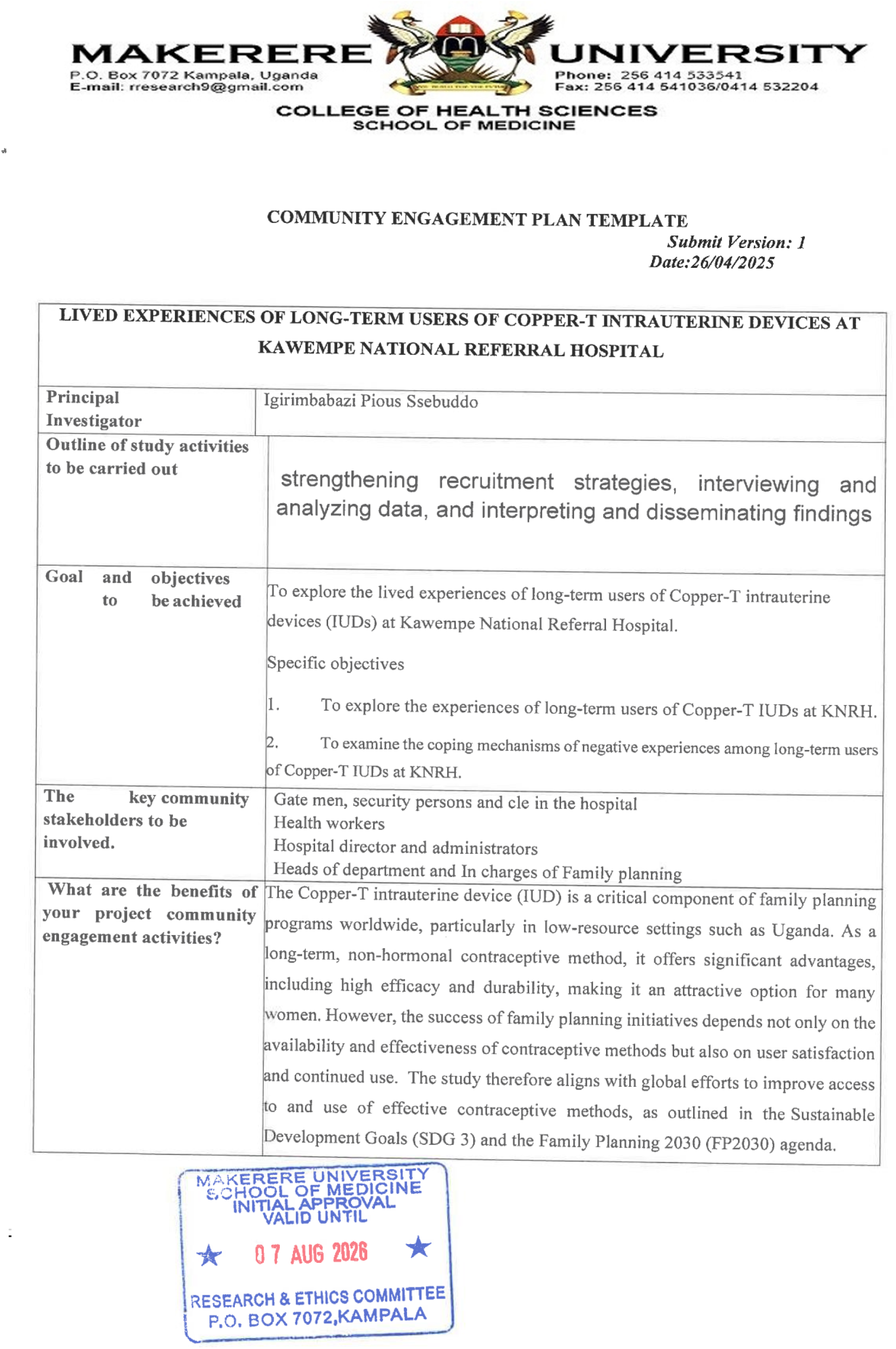
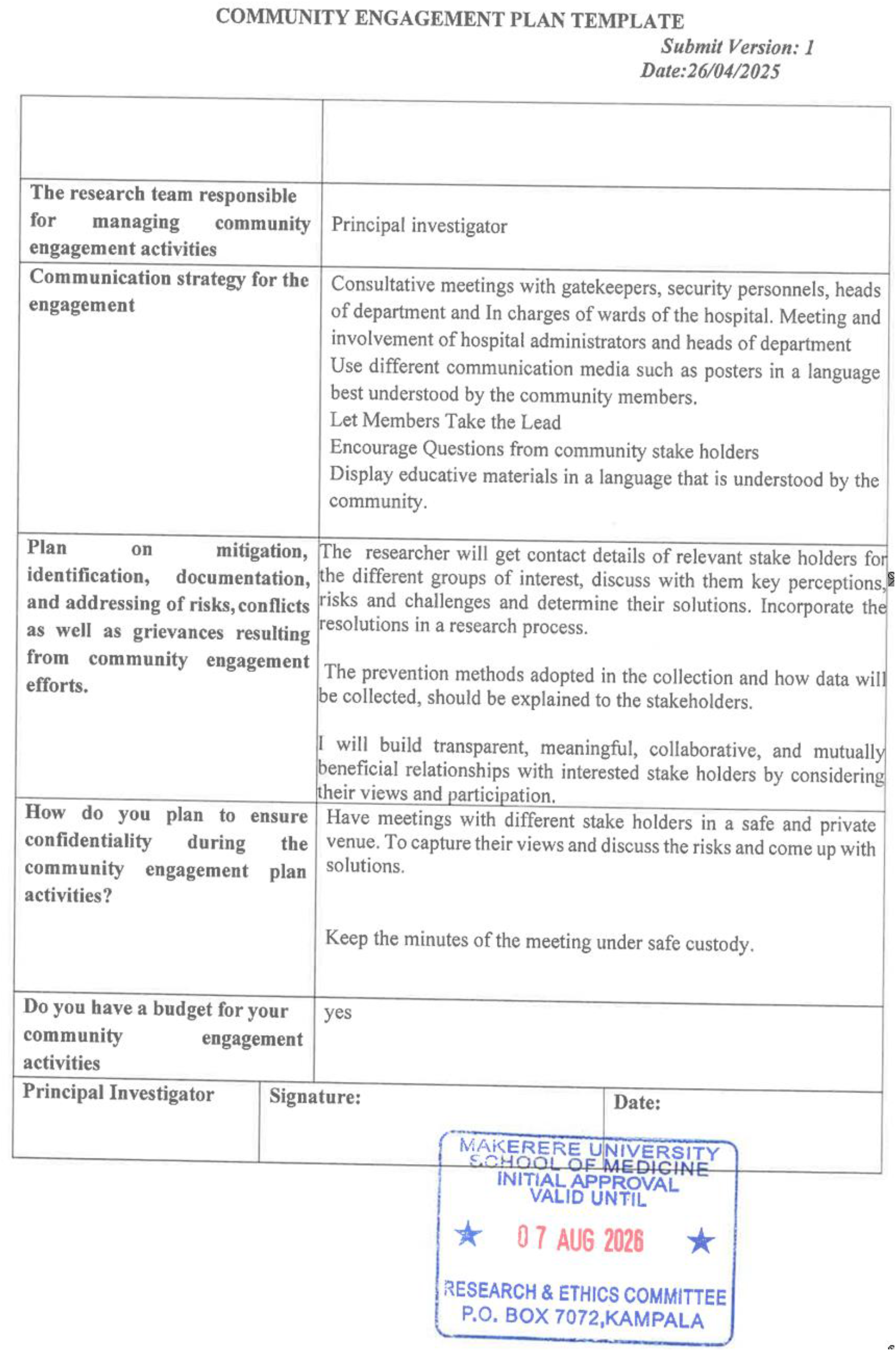

## APPENDIX IV COVID 19 RISK MANAGEMENT

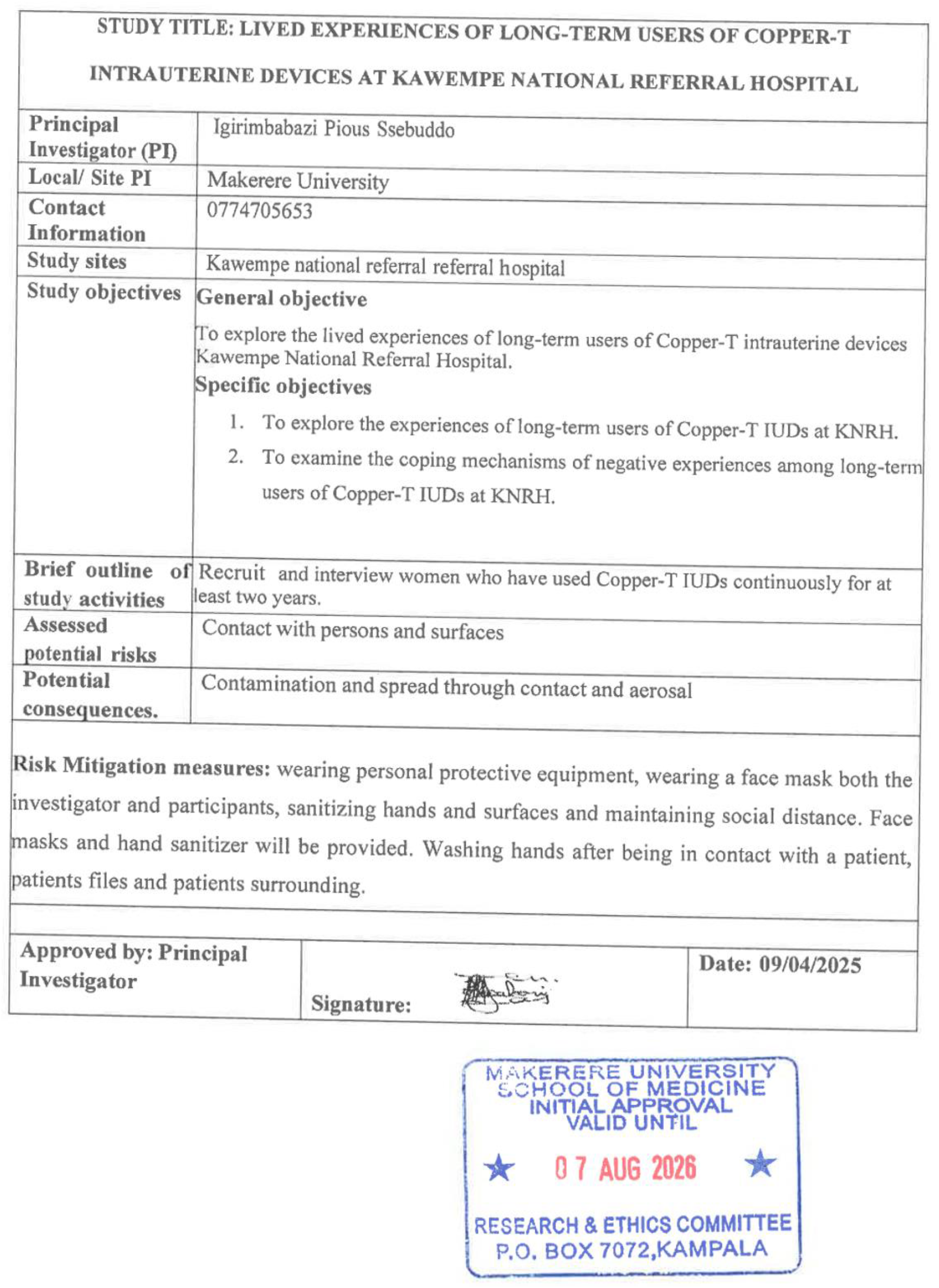

## APPENDIX V SOMREC APPROVAL

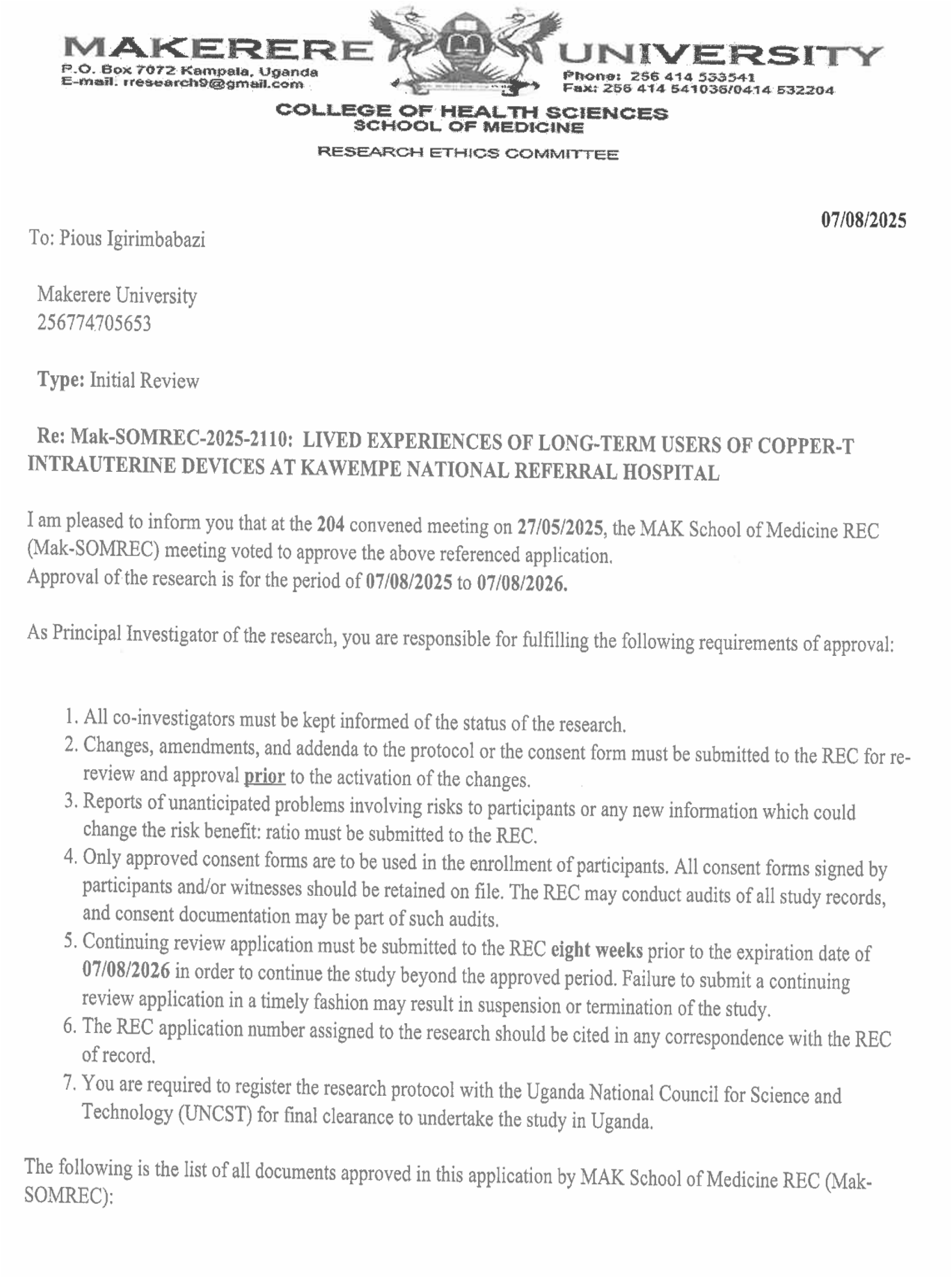
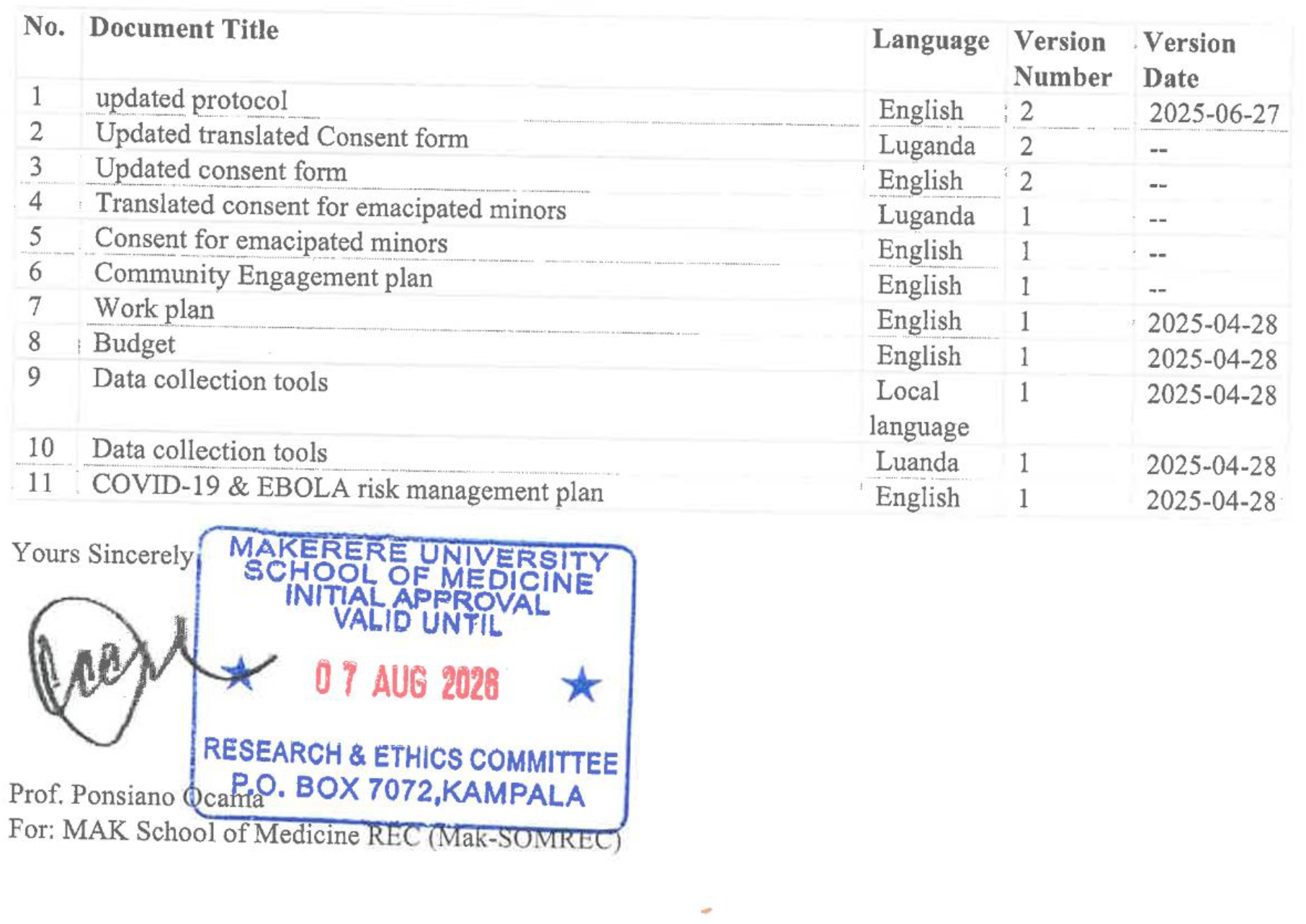

## APPENDIX VI HOSPITAL ADMINISTRATION CLEARANCE

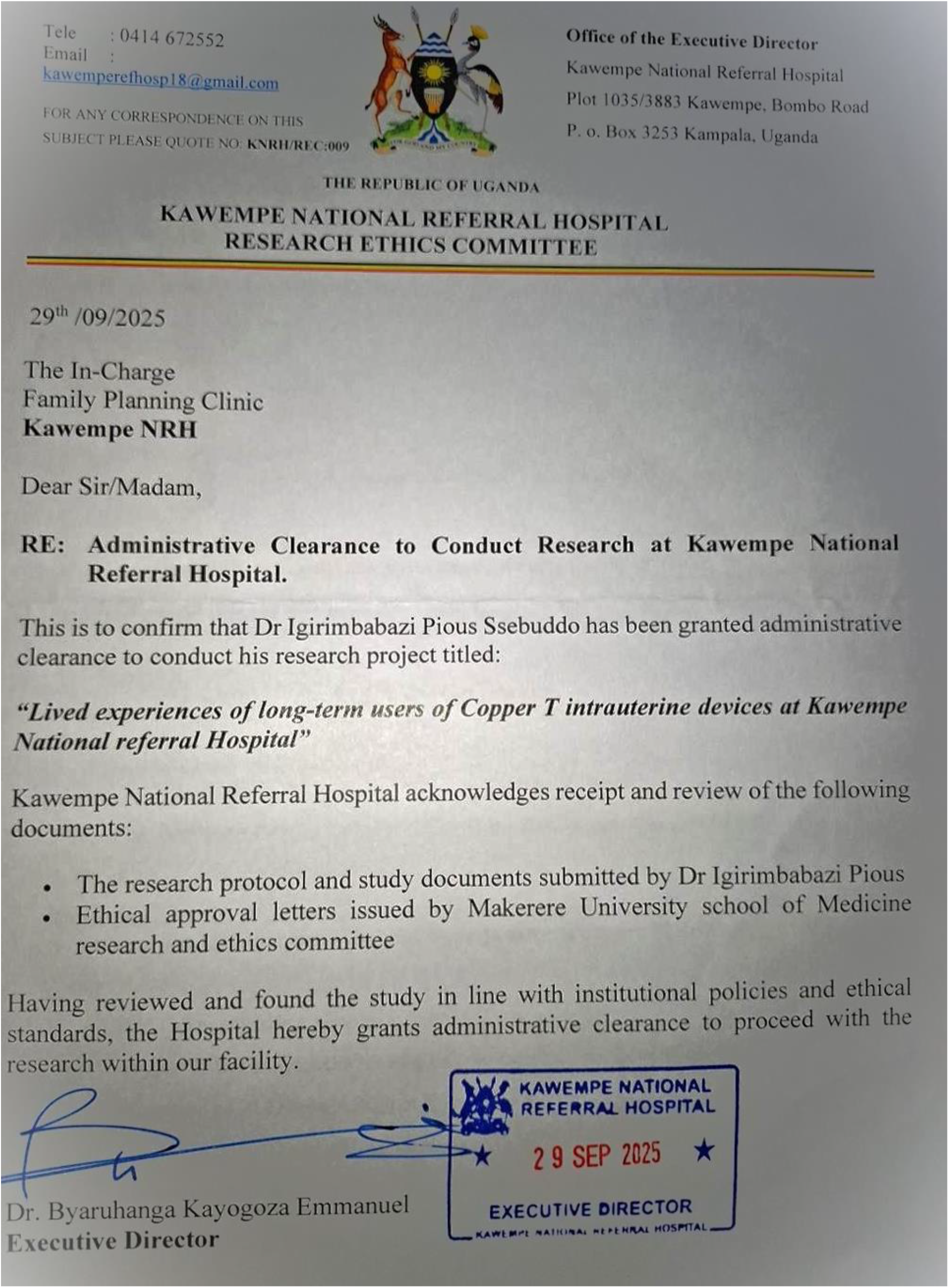

**S1 Checklist. COREQ Checklist — Consolidated Criteria for Reporting Qualitative Research**

Manuscript: Lived Experiences of Long-Term Users of Copper-T Intrauterine Devices at Kawempe National Referral Hospital — A Qualitative Phenomenological Study

Author: Dr. Pious Igirimbabazi Ssebuddo

Supervisors: Dr. Musa Sekikubo, Dr. Imelda Namagembe, Dr. Jolly Beyeza-Kashesya

Reference: Tong A, Sainsbury P, Craig J. Consolidated criteria for reporting qualitative research (COREQ): a 32-item checklist for interviews and focus groups. Int J Qual Health Care. 2007;19(6):349–357.

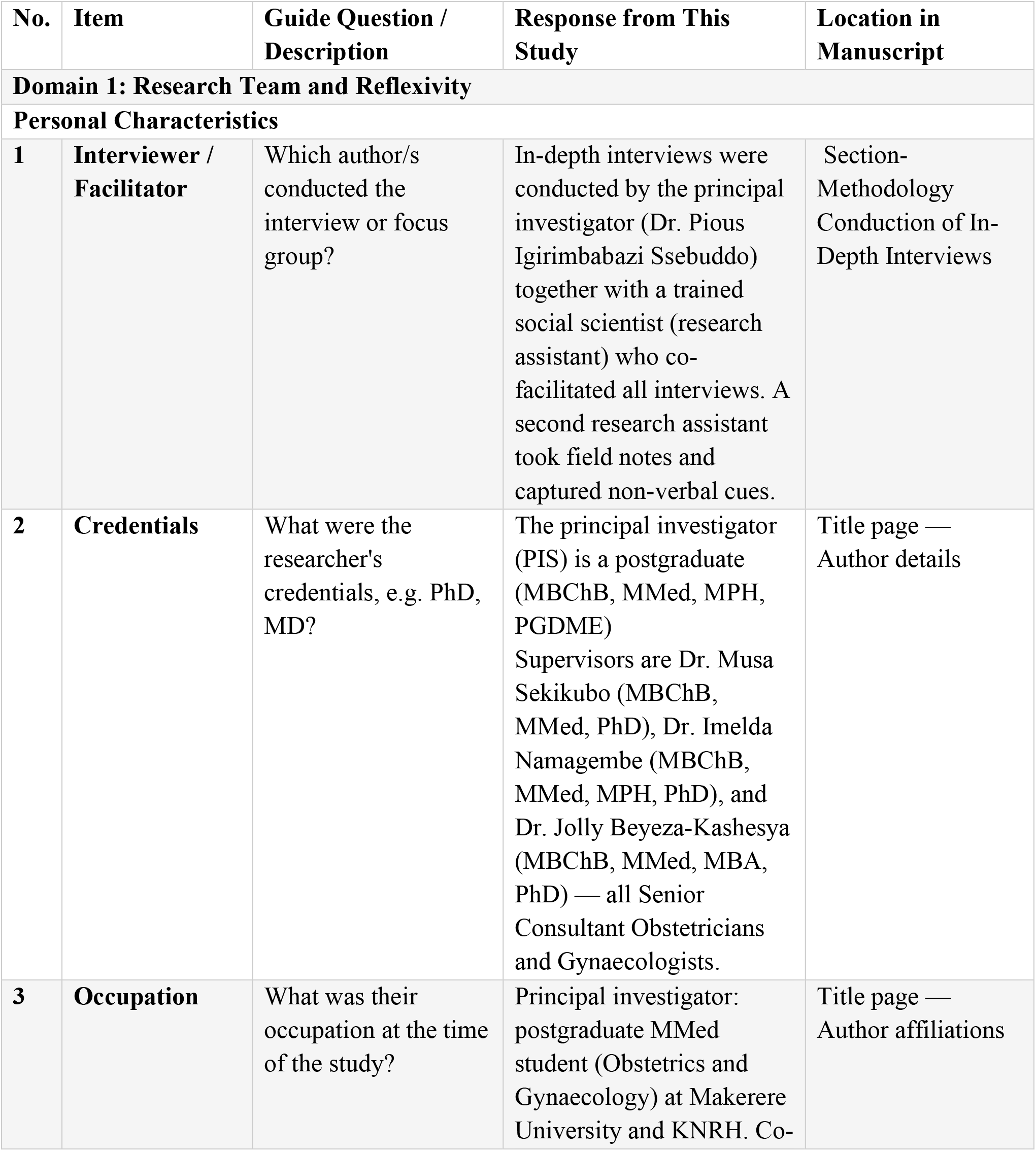
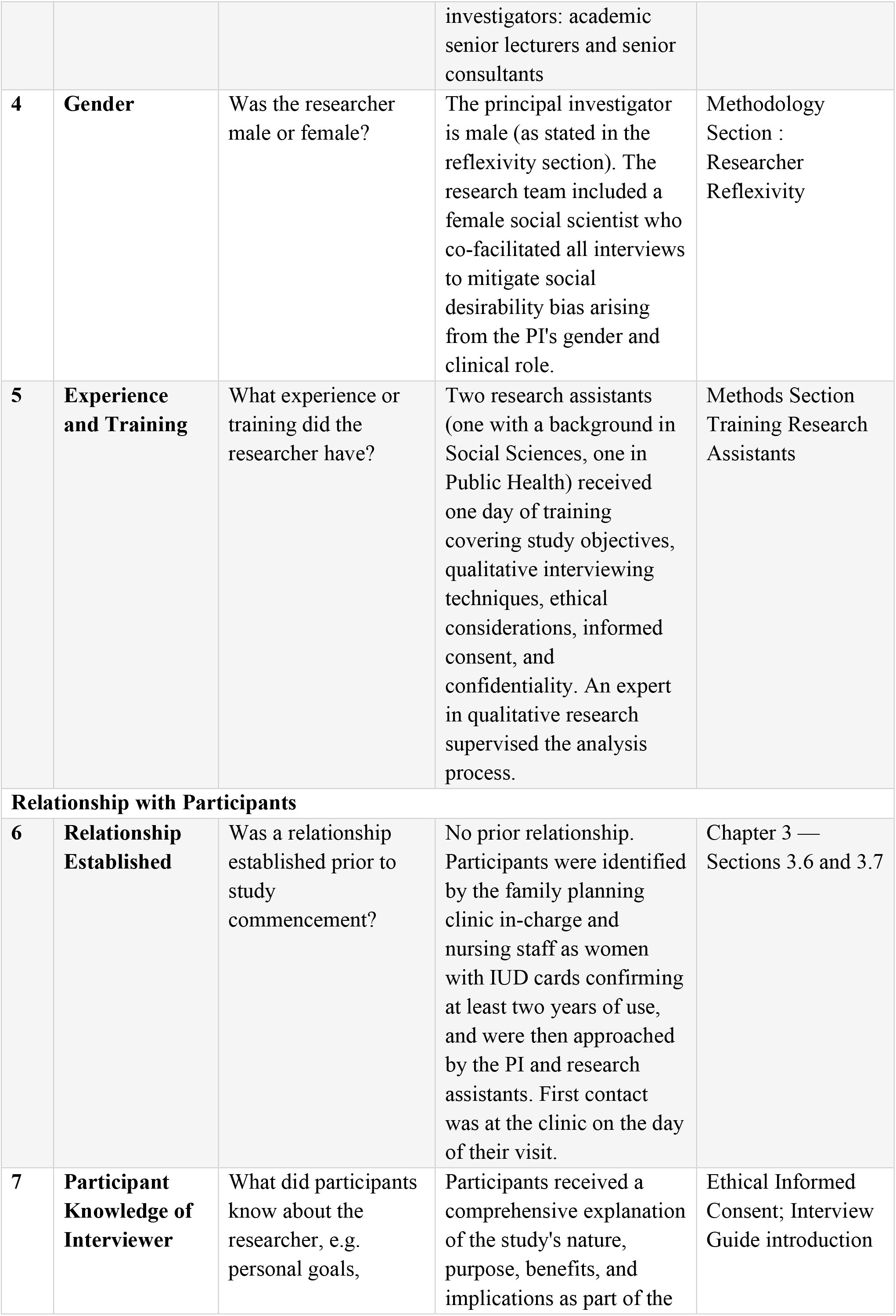
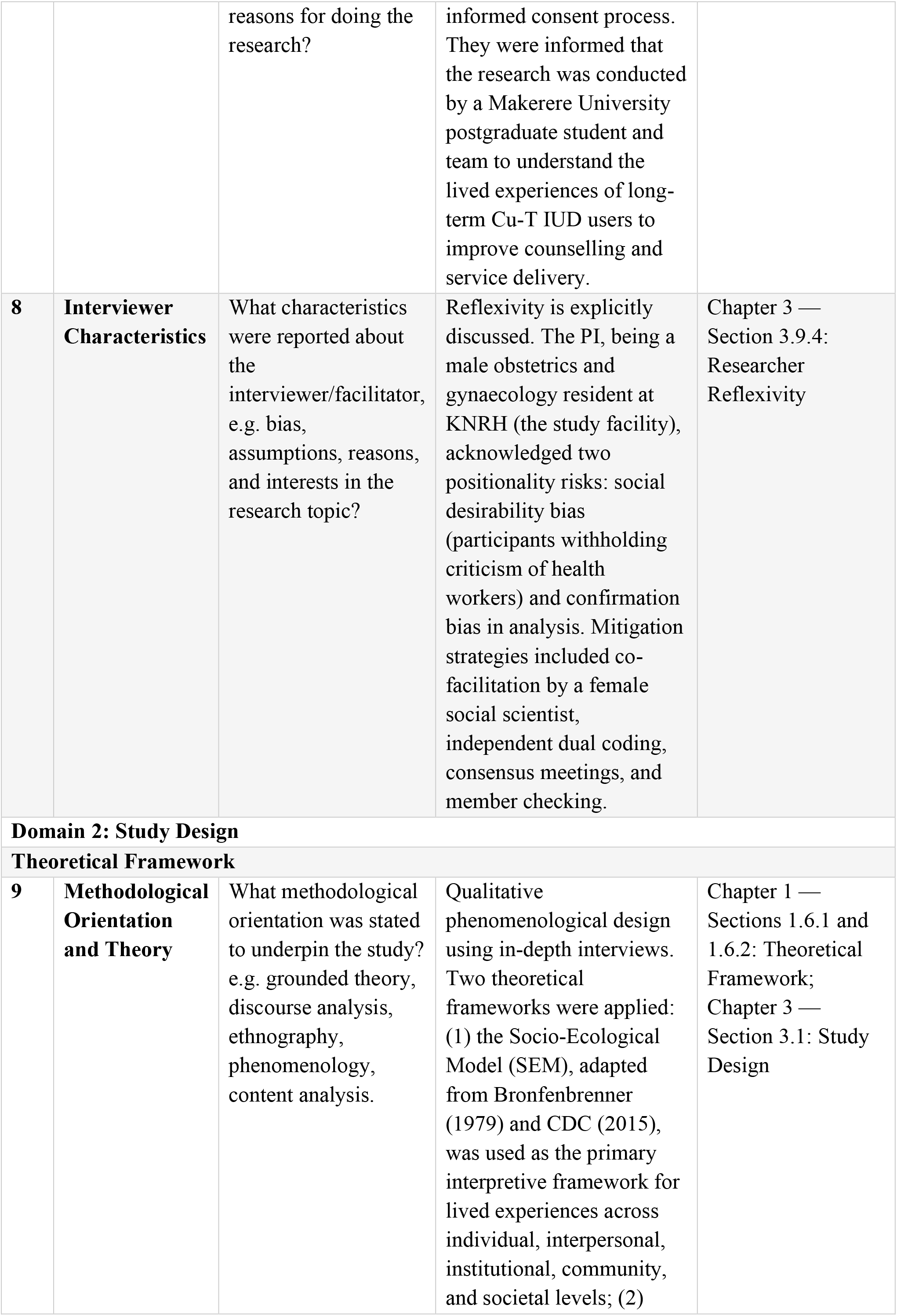
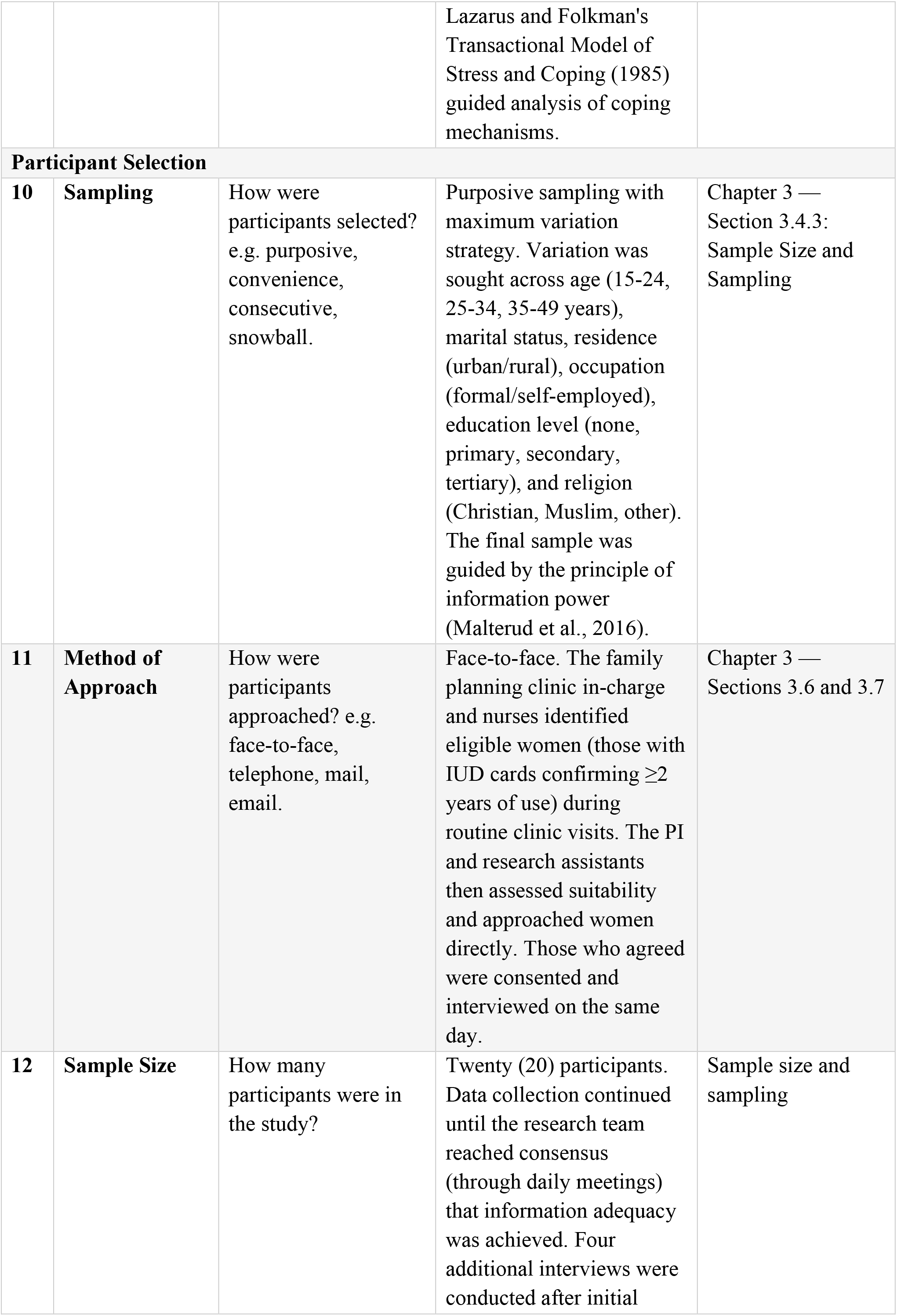
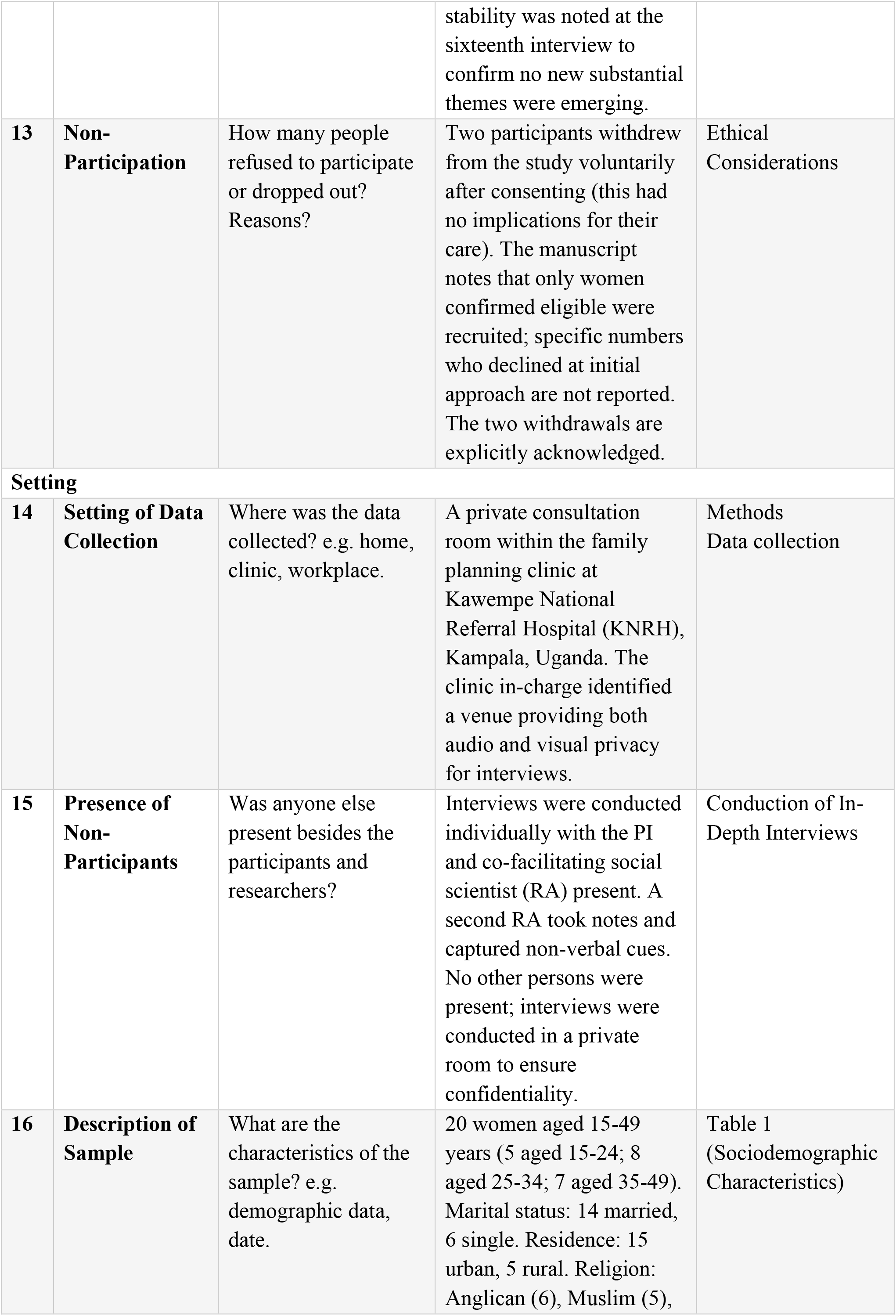
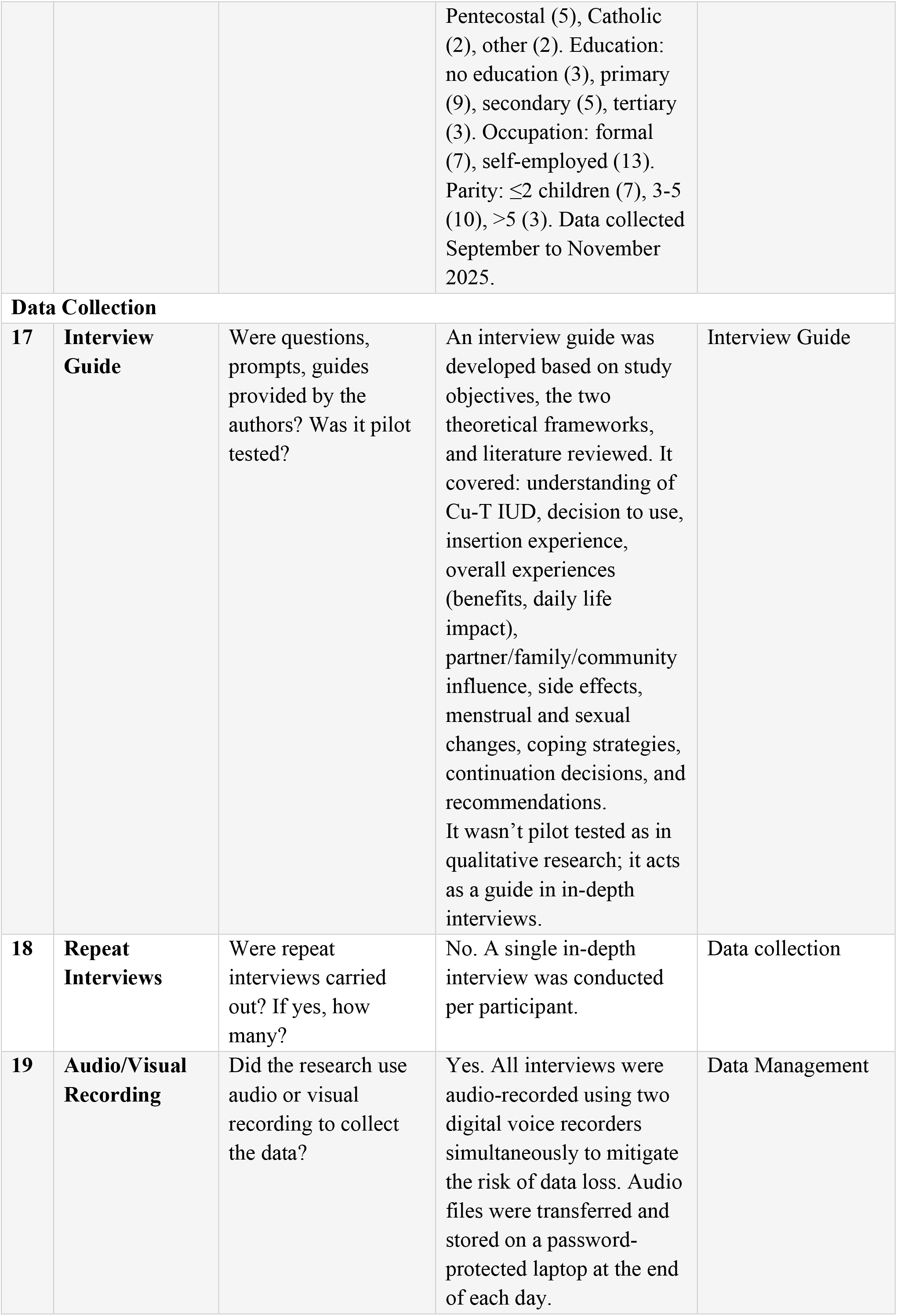
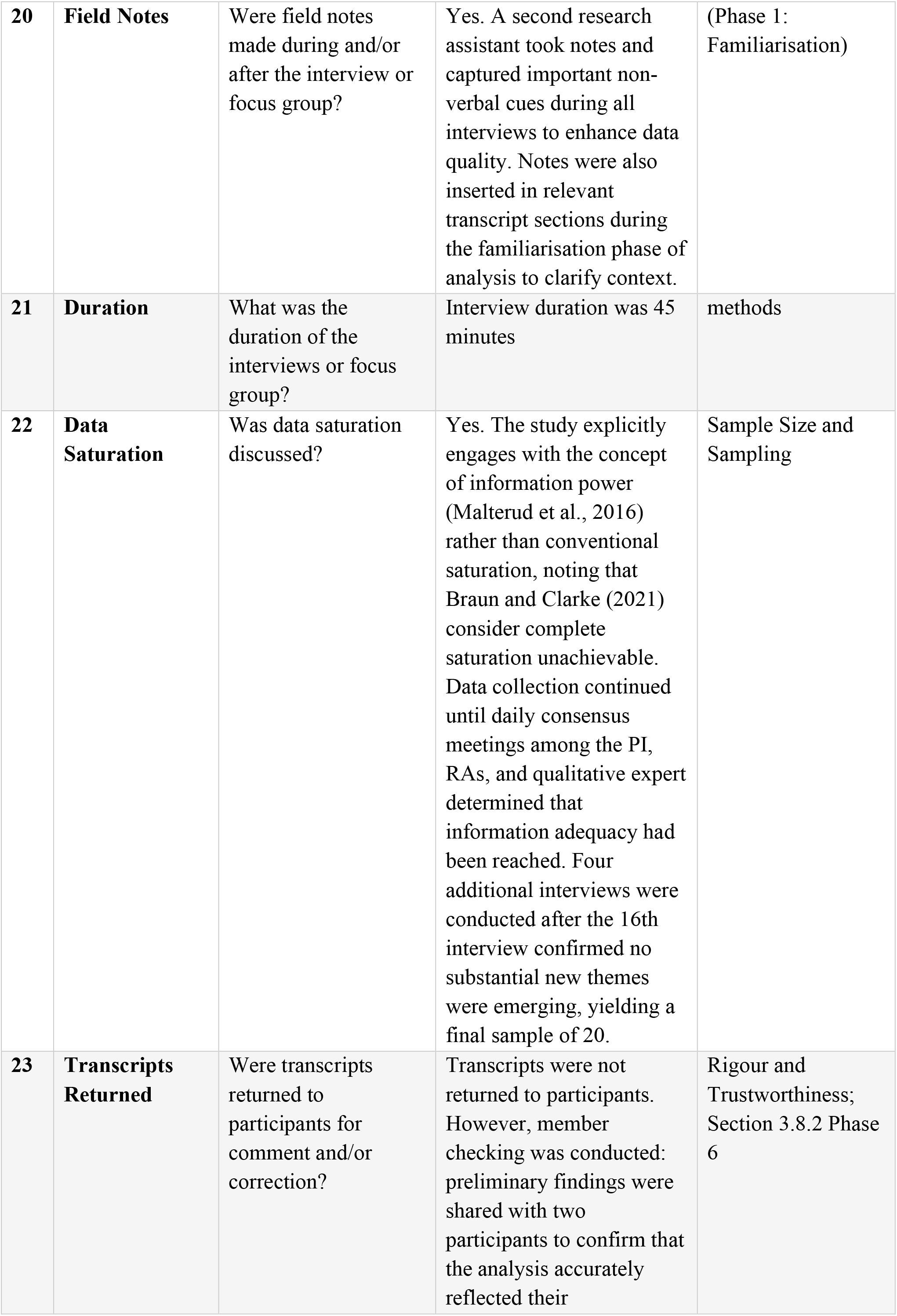
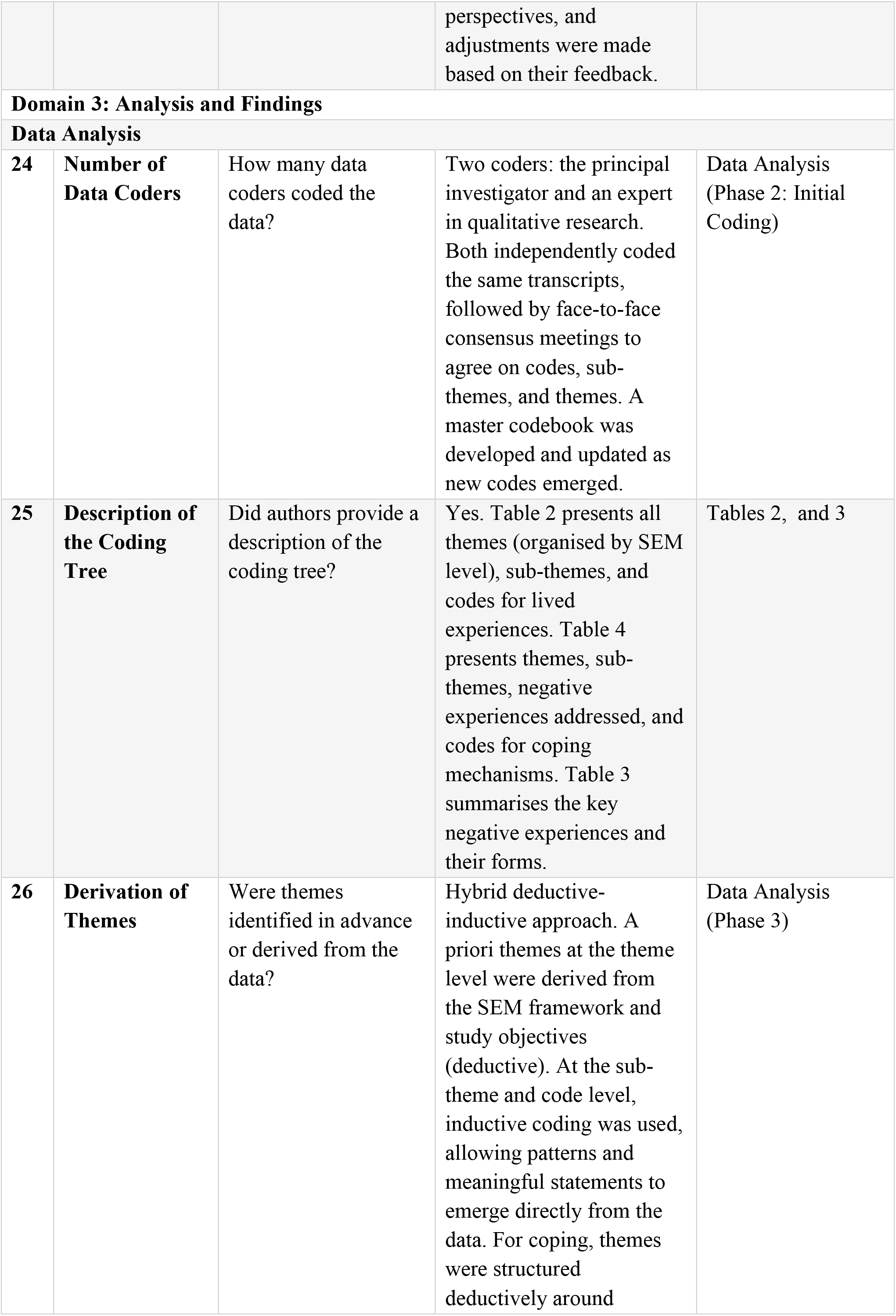
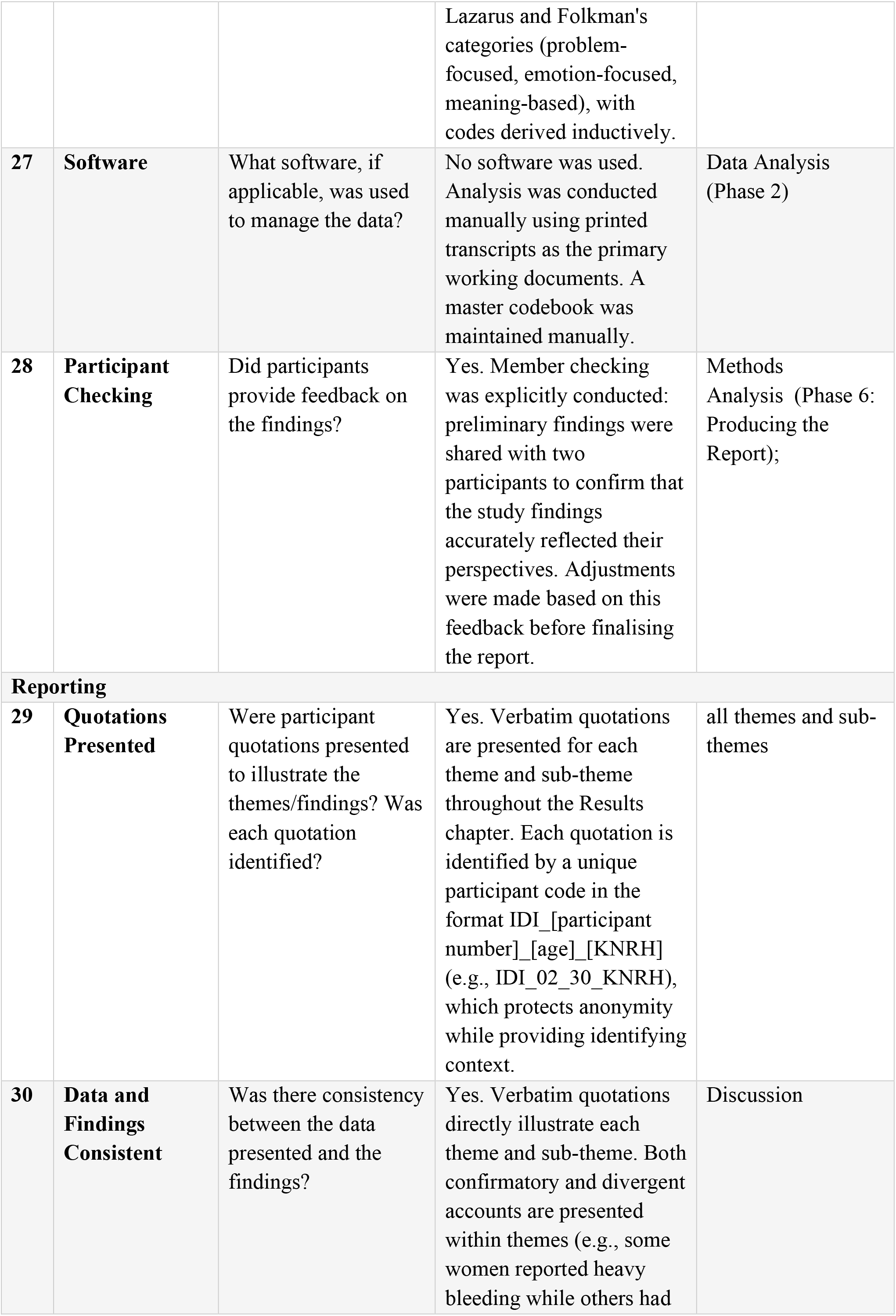
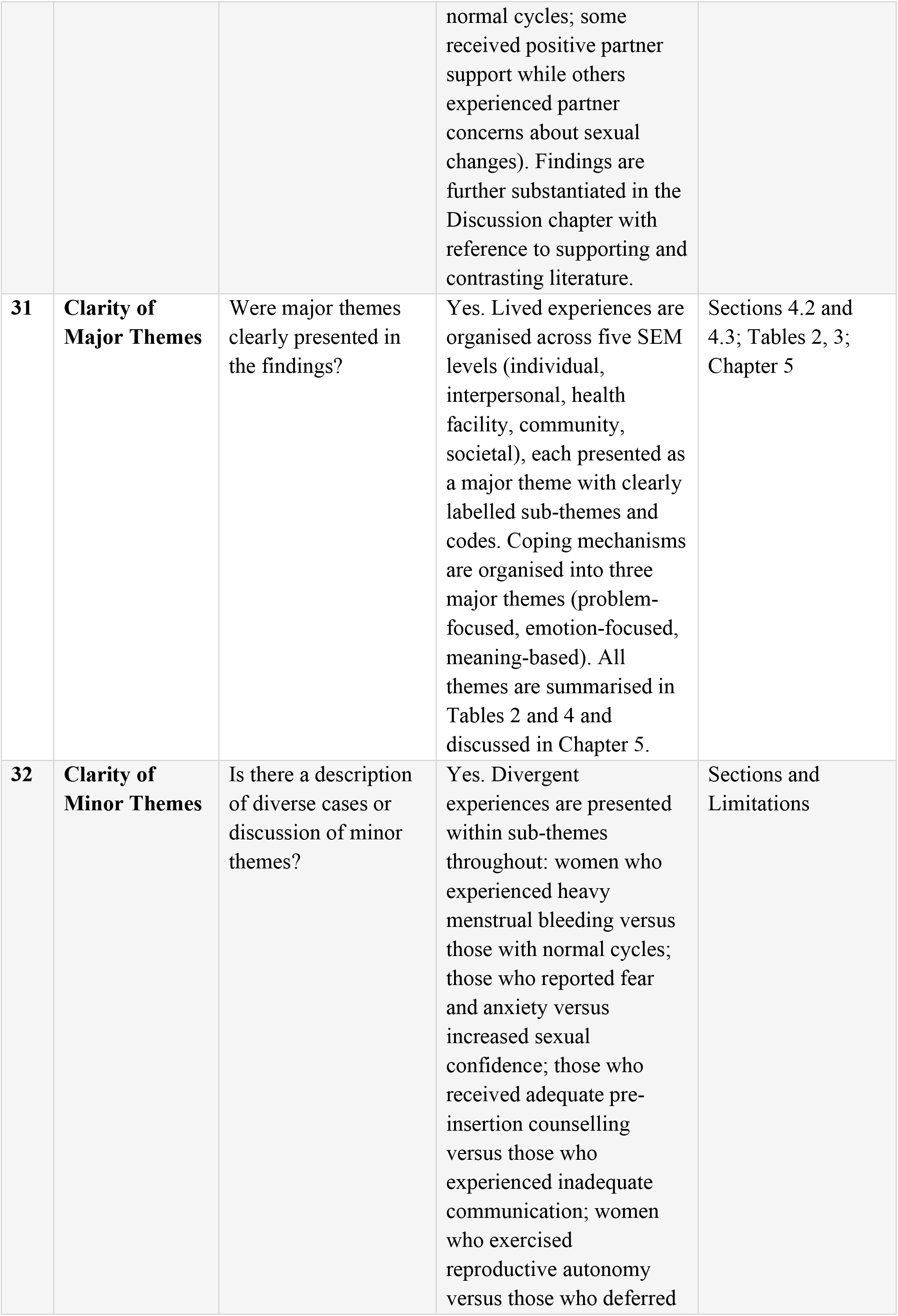
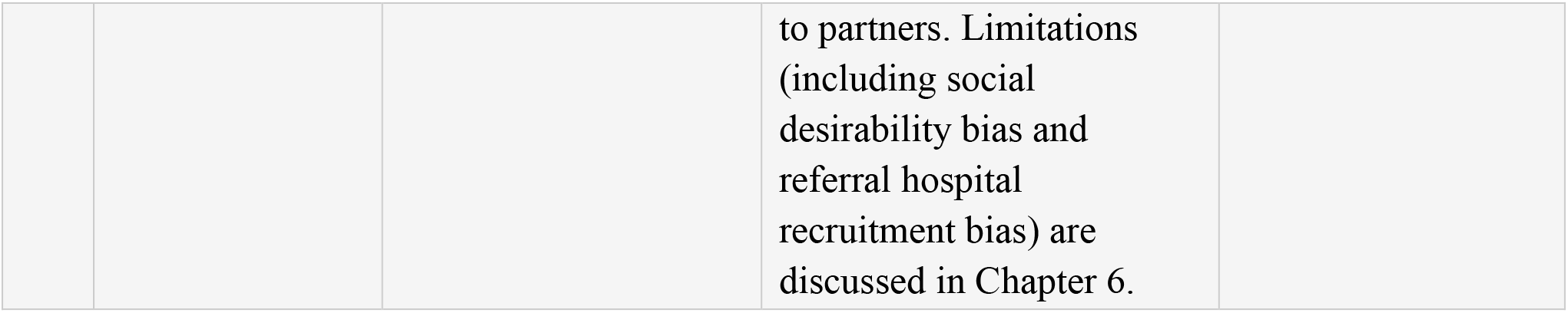

| **No.** | **Item** | **Guide Question / Description** | **Response from This Study** | **Location in Manuscript** |
|---|---|---|---|---|
| **Domain 1: Research Team and Reflexivity** |  |  |  |  |
| **Personal Characteristics** |  |  |  |  |
| **1** | **Interviewer / Facilitator** | Which author/s conducted the interview or focus group? | In-depth interviews were conducted by the principal investigator (Dr. Pious Igirimbabazi Ssebuddo) together with a trained social scientist (research assistant) who co-facilitated all interviews. A second research assistant took field notes and captured non-verbal cues. | Section-Methodology Conduction of In-Depth Interviews |
| **2** | **Credentials** | What were the researcher's credentials, e.g. PhD, MD? | The principal investigator (PIS) is a postgraduate (MBChB, MMed, MPH, PGDME)<br>Supervisors are Dr. Musa Sekikubo (MBChB, MMed, PhD), Dr. Imelda Namagembe (MBChB, MMed, MPH, PhD), and Dr. Jolly Beyeza-Kashesya (MBChB, MMed, MBA, PhD) — all Senior Consultant Obstetricians and Gynaecologists. | Title page — Author details |
| **3** | **Occupation** | What was their occupation at the time of the study? | Principal investigator: postgraduate MMed student (Obstetrics and Gynaecology) at Makerere University and KNRH. Co-investigators: academic senior lecturers and senior consultants | Title page — Author affiliations |
| **4** | **Gender** | Was the researcher male or female? | The principal investigator is male (as stated in the reflexivity section). The research team included a female social scientist who co-facilitated all interviews to mitigate social desirability bias arising from the PI's gender and clinical role. | Methodology Section : Researcher Reflexivity |
| **5** | **Experience and Training** | What experience or training did the researcher have? | Two research assistants (one with a background in Social Sciences, one in Public Health) received one day of training covering study objectives, qualitative interviewing techniques, ethical considerations, informed consent, and confidentiality. An expert in qualitative research supervised the analysis process. | Methods Section Training Research Assistants |
| **Relationship with Participants** |  |  |  |  |
| **6** | **Relationship Established** | Was a relationship established prior to study commencement? | No prior relationship. Participants were identified by the family planning clinic in-charge and nursing staff as women with IUD cards confirming at least two years of use, and were then approached by the PI and research assistants. First contact was at the clinic on the day of their visit. | Chapter 3 — Sections 3.6 and 3.7 |
| **7** | **Participant Knowledge of Interviewer** | What did participants know about the researcher, e.g. personal goals, reasons for doing the research? | Participants received a comprehensive explanation of the study's nature, purpose, benefits, and implications as part of the informed consent process. They were informed that the research was conducted by a Makerere University postgraduate student and team to understand the lived experiences of long-term Cu-T IUD users to improve counselling and service delivery. | Ethical Informed Consent; Interview Guide introduction |
| **8** | **Interviewer Characteristics** | What characteristics were reported about the interviewer/facilitator, e.g. bias, assumptions, reasons, and interests in the research topic? | Reflexivity is explicitly discussed. The PI, being a male obstetrics and gynaecology resident at KNRH (the study facility), acknowledged two positionality risks: social desirability bias (participants withholding criticism of health workers) and confirmation bias in analysis. Mitigation strategies included co-facilitation by a female social scientist, independent dual coding, consensus meetings, and member checking. | Chapter 3 — Section 3.9.4: Researcher Reflexivity |
| **Domain 2: Study Design** |  |  |  |  |
| **Theoretical Framework** |  |  |  |  |
| **9** | **Methodological Orientation and Theory** | What methodological orientation was stated to underpin the study? e.g. grounded theory, discourse analysis, ethnography, phenomenology, content analysis. | Qualitative phenomenological design using in-depth interviews. Two theoretical frameworks were applied: (1) the Socio-Ecological Model (SEM), adapted from Bronfenbrenner (1979) and CDC (2015), was used as the primary interpretive framework for lived experiences across individual, interpersonal, institutional, community, and societal levels; (2) Lazarus and Folkman's Transactional Model of Stress and Coping (1985) guided analysis of coping mechanisms. | Chapter 1 — Sections 1.6.1 and 1.6.2: Theoretical Framework; Chapter 3 — Section 3.1: Study Design |
| **Participant Selection** |  |  |  |  |
| **10** | **Sampling** | How were participants selected? e.g. purposive, convenience, consecutive, snowball. | Purposive sampling with maximum variation strategy. Variation was sought across age (15-24, 25-34, 35-49 years), marital status, residence (urban/rural), occupation (formal/self-employed), education level (none, primary, secondary, tertiary), and religion (Christian, Muslim, other). The final sample was guided by the principle of information power (Malterud et al., 2016). | Chapter 3 — Section 3.4.3: Sample Size and Sampling |
| **11** | **Method of Approach** | How were participants approached? e.g. face-to-face, telephone, mail, email. | Face-to-face. The family planning clinic in-charge and nurses identified eligible women (those with IUD cards confirming ≥2 years of use) during routine clinic visits. The PI and research assistants then assessed suitability and approached women directly. Those who agreed were consented and interviewed on the same day. | Chapter 3 — Sections 3.6 and 3.7 |
| **12** | **Sample Size** | How many participants were in the study? | Twenty (20) participants. Data collection continued until the research team reached consensus (through daily meetings) that information adequacy was achieved. Four additional interviews were conducted after initial stability was noted at the sixteenth interview to confirm no new substantial themes were emerging. | Sample size and sampling |
| **13** | **Non-Participation** | How many people refused to participate or dropped out? Reasons? | Two participants withdrew from the study voluntarily after consenting (this had no implications for their care). The manuscript notes that only women confirmed eligible were recruited; specific numbers who declined at initial approach are not reported. The two withdrawals are explicitly acknowledged. | Ethical Considerations |
| **Setting** |  |  |  |  |
| **14** | **Setting of Data Collection** | Where was the data collected? e.g. home, clinic, workplace. | A private consultation room within the family planning clinic at Kawempe National Referral Hospital (KNRH), Kampala, Uganda. The clinic in-charge identified a venue providing both audio and visual privacy for interviews. | Methods<br>Data collection |
| **15** | **Presence of Non-Participants** | Was anyone else present besides the participants and researchers? | Interviews were conducted individually with the PI and co-facilitating social scientist (RA) present. A second RA took notes and captured non-verbal cues. No other persons were present; interviews were conducted in a private room to ensure confidentiality. | Conduction of In-Depth Interviews |
| **16** | **Description of Sample** | What are the characteristics of the sample? e.g. demographic data, date. | 20 women aged 15-49 years (5 aged 15-24; 8 aged 25-34; 7 aged 35-49). Marital status: 14 married, 6 single. Residence: 15 urban, 5 rural. Religion: Anglican (6), Muslim (5), Pentecostal (5), Catholic (2), other (2). Education: no education (3), primary (9), secondary (5), tertiary (3). Occupation: formal (7), self-employed (13). Parity: ≤2 children (7), 3-5 (10), >5 (3). Data collected September to November 2025. | Table 1 (Sociodemographic Characteristics) |
| **Data Collection** |  |  |  |  |
| **17** | **Interview Guide** | Were questions, prompts, guides provided by the authors? Was it pilot tested? | An interview guide was developed based on study objectives, the two theoretical frameworks, and literature reviewed. It covered: understanding of Cu-T IUD, decision to use, insertion experience, overall experiences (benefits, daily life impact), partner/family/community influence, side effects, menstrual and sexual changes, coping strategies, continuation decisions, and recommendations.<br>It wasn’t pilot tested as in qualitative research; it acts as a guide in in-depth interviews. | Interview Guide |
| **18** | **Repeat Interviews** | Were repeat interviews carried out? If yes, how many? | No. A single in-depth interview was conducted per participant. | Data collection |
| **19** | **Audio/Visual Recording** | Did the research use audio or visual recording to collect the data? | Yes. All interviews were audio-recorded using two digital voice recorders simultaneously to mitigate the risk of data loss. Audio files were transferred and stored on a password-protected laptop at the end of each day. | Data Management |
| **20** | **Field Notes** | Were field notes made during and/or after the interview or focus group? | Yes. A second research assistant took notes and captured important non-verbal cues during all interviews to enhance data quality. Notes were also inserted in relevant transcript sections during the familiarisation phase of analysis to clarify context. | (Phase 1: Familiarisation) |
| **21** | **Duration** | What was the duration of the interviews or focus group? | Interview duration was 45 minutes | methods |
| **22** | **Data Saturation** | Was data saturation discussed? | Yes. The study explicitly engages with the concept of information power (Malterud et al., 2016) rather than conventional saturation, noting that Braun and Clarke (2021) consider complete saturation unachievable. Data collection continued until daily consensus meetings among the PI, RAs, and qualitative expert determined that information adequacy had been reached. Four additional interviews were conducted after the 16th interview confirmed no substantial new themes were emerging, yielding a final sample of 20. | Sample Size and Sampling |
| **23** | **Transcripts Returned** | Were transcripts returned to participants for comment and/or correction? | Transcripts were not returned to participants. However, member checking was conducted: preliminary findings were shared with two participants to confirm that the analysis accurately reflected their perspectives, and adjustments were made based on their feedback. | Rigour and Trustworthiness; Section 3.8.2 Phase 6 |
| **Domain 3: Analysis and Findings** |  |  |  |  |
| **Data Analysis** |  |  |  |  |
| **24** | **Number of Data Coders** | How many data coders coded the data? | Two coders: the principal investigator and an expert in qualitative research. Both independently coded the same transcripts, followed by face-to-face consensus meetings to agree on codes, sub-themes, and themes. A master codebook was developed and updated as new codes emerged. | Data Analysis (Phase 2: Initial Coding) |
| **25** | **Description of the Coding Tree** | Did authors provide a description of the coding tree? | Yes. Table 2 presents all themes (organised by SEM level), sub-themes, and codes for lived experiences. Table 4 presents themes, sub-themes, negative experiences addressed, and codes for coping mechanisms. Table 3 summarises the key negative experiences and their forms. | Tables 2, and 3 |
| **26** | **Derivation of Themes** | Were themes identified in advance or derived from the data? | Hybrid deductive-inductive approach. A priori themes at the theme level were derived from the SEM framework and study objectives (deductive). At the sub-theme and code level, inductive coding was used, allowing patterns and meaningful statements to emerge directly from the data. For coping, themes were structured deductively around Lazarus and Folkman's categories (problem-focused, emotion-focused, meaning-based), with codes derived inductively. | Data Analysis (Phase 3) |
| **27** | **Software** | What software, if applicable, was used to manage the data? | No software was used. Analysis was conducted manually using printed transcripts as the primary working documents. A master codebook was maintained manually. | Data Analysis (Phase 2) |
| **28** | **Participant Checking** | Did participants provide feedback on the findings? | Yes. Member checking was explicitly conducted: preliminary findings were shared with two participants to confirm that the study findings accurately reflected their perspectives. Adjustments were made based on this feedback before finalising the report. | Methods<br>Analysis (Phase 6: Producing the Report); |
| **Reporting** |  |  |  |  |
| **29** | **Quotations Presented** | Were participant quotations presented to illustrate the themes/findings? Was each quotation identified? | Yes. Verbatim quotations are presented for each theme and sub-theme throughout the Results chapter. Each quotation is identified by a unique participant code in the format IDI_[participant number]_[age]_[KNRH] (e.g., IDI_02_30_KNRH), which protects anonymity while providing identifying context. | all themes and sub-themes |
| **30** | **Data and Findings Consistent** | Was there consistency between the data presented and the findings? | Yes. Verbatim quotations directly illustrate each theme and sub-theme. Both confirmatory and divergent accounts are presented within themes (e.g., some women reported heavy bleeding while others had normal cycles; some received positive partner support while others experienced partner concerns about sexual changes). Findings are further substantiated in the Discussion chapter with reference to supporting and contrasting literature. | Discussion |
| **31** | **Clarity of Major Themes** | Were major themes clearly presented in the findings? | Yes. Lived experiences are organised across five SEM levels (individual, interpersonal, health facility, community, societal), each presented as a major theme with clearly labelled sub-themes and codes. Coping mechanisms are organised into three major themes (problem-focused, emotion-focused, meaning-based). All themes are summarised in Tables 2 and 4 and discussed in Chapter 5. | Sections 4.2 and 4.3; Tables 2, 3; Chapter 5 |
| **32** | **Clarity of Minor Themes** | Is there a description of diverse cases or discussion of minor themes? | Yes. Divergent experiences are presented within sub-themes throughout: women who experienced heavy menstrual bleeding versus those with normal cycles; those who reported fear and anxiety versus increased sexual confidence; those who received adequate pre-insertion counselling versus those who experienced inadequate communication; women who exercised reproductive autonomy versus those who deferred to partners. Limitations (including social desirability bias and referral hospital recruitment bias) are discussed in Chapter 6. | Sections and Limitations |

## References

1. United Nations Department of Economic and Social Affairs. World family planning 2022: meeting the changing needs for family planning: contraceptive use by age and method. 2022.

2. Ayogu ME, Omonua KI, Ayogu MC. A 10-year experience with copper T intrauterine device (IUD) in Abuja, Nigeria. Niger J Med. 2019;28(2):161–7.

3. Phiri M, Odimegwu C, Kalinda C. Unmet need for family planning among married women in sub-Saharan Africa: a meta-analysis of DHS data (1995-2020. Contraception and reproductive medicine. 2023;8(1).

4. Teshale AB. Factors associated with unmet need for family planning in sub-Saharan Africa: a multilevel multinomial logistic regression analysis. PLoS One. 2022;17(2).

5. Uganda Demographic and Health Survey 2022 Kampala: UBOS; 2022 [updated 2022. Available from: https://www.unicef.org/uganda/media/16731/file/UDHS-2022-Report.pdf.pdf.

6. Family planning: a global handbook for providers Geneva: WHO; 2022 [updated 2022. Available from: https://cdn.who.int/media/docs/default-source/reproductive-health/contraception-family-planning/family-planning-a-global-handbook-for-providers-2022.pdf.

7. Lanzola EL, Auber M, Ketvertis K. Intrauterine device placement and removal. StatPearls [Internet. Treasure Island: StatPearls Publishing; 2025.

8. Bameka A, Kakaire O, Kaye DK, Namusoke F. Early discontinuation of long-acting reversible contraceptives and associated factors among women discontinuing long-acting reversible contraceptives at a national referral hospital, Kampala-Uganda; a cross-sectional study. Contracept Reprod Med. 2023;8(1).

9. Boadu I. Coverage and determinants of modern contraceptive use in sub-Saharan Africa: further analysis of demographic and health surveys. Reprod Health. 2022;19(1).

10. Jean de Dieu H, Lambert M. Predictors of contraceptive discontinuation in Rwanda: evidence from demographic and health survey 2019–2020. Contraception and Reproductive Medicine. 2024;9(1):19.

11. Kungu W, Agwanda A, Khasakhala A. Prevalence of and factors associated with contraceptive discontinuation in Kenya. African journal of primary health care & family medicine. 2022;14(1):2992.

12. Alo OD, Daini BO, Omisile OK, Ubah EJ, Adelusi OE, Idoko-Asuelimhen O. Factors influencing the use of modern contraceptive in Nigeria: a multilevel logistic analysis using linked data from performance monitoring and accountability 2020. BMC Womens Health. 2020;20(1).

13. Sanders JN, Adkins DE, Kaur S, Storck K, Gawron LM, Turok DK. Bleeding, cramping, and satisfaction among new copper IUD users: a prospective study. PLoS One. 2018;13(11).

14. Buser JM, Pebolo PF, August E, Rana GK, Gray R, Jacobson-Davies FE. Scoping review of qualitative studies on family planning in Uganda. PLOS Glob Public Health. 2024;4(7).

15. Neubauer BE, Witkop CT, Varpio L. How phenomenology can help us learn from the experiences of others. Perspect Med Educ. 2019;8(2):90–7.

16. UNICEF. Key practice: spacing between pregnancies [Internet Kampala: UNICEF; 2020 [updated 2020. Available from: https://www.unicef.org/uganda/key-practice-spacing-between-pregnancies.

17. Creswell JW, Poth CN. Qualitative inquiry and research design: choosing among five approaches. 4th ed. Thousand Oaks: SAGE Publications; 2018 2018.

18. Malterud K, Siersma VD, Guassora AD. Sample size in qualitative interview studies: guided by information power. Qual Health Res. 2016;26(13):1753–60.

19. Braun V, Clarke V. To saturate or not to saturate? Questioning data saturation as a useful concept for thematic analysis and sample-size rationales. Qualitative research in sport, exercise and health. 2021;13(2):201–16.

20. Selected practice recommendations for contraceptive use Geneva: WHO; 2024 [updated 2024. 4th [Available from: https://www.who.int/publications/i/item/9789240115606.

21. Nakate K, Bantia M, Naggulu IP, Nalubega JF. Factors influencing utilization of intrauterine contraceptive devices among women of reproductive age at Entebbe Regional Referral Hospital, Wakiso District: a cross-sectional study. Stud J Health Res Afr. 2025;6(3).

22. Hubacher D, Kavanaugh M. Historical record-setting trends in IUD use in the United States. Contraception. 2018;98(6):467–70.

23. Blackstone N, Sr., U I, J. Factors influencing contraceptive use in sub-Saharan Africa: a systematic review. Int Q Community Health Educ. 2017;37(2):79–91.

24. Brandi K, Fuentes L. The history of tiered-effectiveness contraceptive counseling and the importance of patient-centered family planning care. American journal of obstetrics and gynecology. 2020;222(4):S873–S7.

25. Rodriguez MB, Dias S, Jordan V, Lethaby A, Lensen SF, Wise MR, et al. Interventions for heavy menstrual bleeding; overview of Cochrane reviews and network meta-analysis. Cochrane Database of Systematic Reviews. 2022(5).

26. Lesko J. How to place an IUD with minimal patient discomfort: Managing your patient’s pain during IUD placement may make it a more comfortable and satisfying experience for both of you. OBG Management. 2023;35(2):19–26.

27. Ehusani R. Stress Coping Mechanisms in the Lord’s Prayer: A Content Analysis.

28. Chidebe RCW, Osayi A, Torode JS. The Global Fund, Cervical Cancer, and HPV infections: what can low- and middle-income countries do to accelerate progress by 2030? eClinicalMedicine. 2025;81.

